# A Model for Understanding the Causes and Consequences of Walking Impairments

**DOI:** 10.1101/2022.06.17.22276552

**Authors:** Michael H. Schwartz, Katherine M. Steele, Andrew J. Ries, Andrew G. Georgiadis, Bruce A. MacWilliams

**Author notes:** These authors contributed equally to this work. These authors contributed equally to this work (Katherine M. Steele, Andrew J. Ries, Andrew G. Georgiadis, Bruce A. MacWilliams).

## Abstract

Walking is an important skill with positive impacts on health, function, and well-being. Many disorders impair walking and its positive impacts through a variety of complex and interrelated mechanisms. Any attempt to understand walking impairments, or the effects of interventions intended to treat these impairments, must respect this complexity. Therefore, our main objectives in conducting this study were to (1) propose a comprehensive model for quantifying the causes and consequences of walking impairments and (2) demonstrate the potential utility of the model for supporting clinical care and addressing basic scientific questions related to walking.

To achieve these goals, we introduced a model, described by a directed acyclic graph, consisting of 10 nodes and 23 primary causal paths. We gave detailed descriptions of each node and path based on domain knowledge. We then demonstrated the model’s utility using a large sample of gait data (N = 9504) acquired as part of routine care at a regional referral center. We analyzed five relevant examples that involved many of the model’s nodes and paths. We computed causal effect magnitudes as Shapley values and displayed the overall importance of variables (mean absolute Shapley value), the variation of Shapley values with respect to underlying variables, and Shapley values for individual observations (case studies).

We showed that the model was plausible, captured some well-known cause-effect relationships, provided new insights into others, and generated novel hypotheses requiring further testing through simulation or experiment. To aid in transparency, reproducibility, and future enhancements we have included an extensively commented Rmarkdown file and a deidentified data set.

## 1 Introduction

### 1.1 Motivation and Purpose

Walking is important. It is the fundamental mode of human movement, contributes to independence, plays a role in activity, participation, and social relations, and promotes health. Walking impairments arise from medical conditions like osteoarthritis, cerebral palsy, stroke, obesity, traumatic injuries, and many others. These impairments are treated with a variety of approaches, such as physical therapy, orthoses, oral and injected pharmaceutical agents, orthopedic surgery, and neurosurgery. To improve or correct walking impairments and their sequelae it is necessary to (1) identify the cause of the impairment, (2) have access to treatments that impact the identified cause, and (3) skillfully deliver the proper dose of the treatment.

Consider a patient with a severely osteoarthritic hip. The patient has a slow, unstable, Trendelenburg gait, has trouble conducting activities of daily living, has stopped participating in leisure activities, and reports that these problems lower their independence and happiness. The causal path from the osteoarthritic hip to the impairments is clear, a well-established and effective treatment (total hip arthroplasty) is available, and the dose (one hip) is not in question. As a result, a total hip arthroplasty for this patient is likely to result in large predictable improvements to walking, mobility, and quality-of-life (1).

Now consider a patient with cerebral palsy (CP) who has spasticity, poor motor control, weakness, muscle contractures, bony torsions, and unstable feet. The patient presents with comparable gait, mobility, and quality-of-life complaints as the osteoarthritic patient. Which of the patient’s underlying orthopedic and neurological problems are the cause of their impairments? What treatments are available for these underlying causes? What dose of correction is appropriate?

Unfortunately, treatment of impairments arising from CP result in modest and unpredictable outcomes that have been stagnant for at least the last several decades (2). We believe that the difficulty in identifying causes of gait impairments is an important contributor to this unsatisfactory situation.

In this study we will propose a comprehensive model for quantifying causes and consequences of walking impairments and demonstrate the potential utility of the model for supporting clinical care decisions and addressing scientific questions.

### 1.2 Causal Inference

Causal inference has been carefully and succinctly defined many times, including for the International Encyclopedia of the Social and Behavioral Sciences (Second Edition). Quoting extensively below [with adaptations for our application to walking impairments in brackets]:

> *“Causal inference refers to an intellectual discipline that considers the assumptions, study designs, and estimation strategies that allow researchers to draw causal conclusions based on data. … The dominant perspective on causal inference in statistics has philosophical underpinnings that rely on consideration of counterfactual states. In particular, it considers the outcomes that could manifest given exposure to each of a set of [causal factors]. Causal effects are defined as comparisons between these ‘potential outcomes.’ For instance, the causal effect of [femoral anteversion] on [in-toeing] would be defined as a comparison of the [in-toeing] that would be measured [with one level of femoral anteversion] with the [in-toeing] that would be measured [with a different magnitude level of femoral anteversion]. The challenge for causal inference is that we are not generally able to observe both of these states: at the point in time when we are measuring the [in-toeing], each individual … has [only one level of femoral anteversion]”* (*3*).

We emphasize that the key difficulty for causal inference is that we can never simultaneously observe the actual and counterfactual states (e.g., the same limb with two different levels of anteversion). As a result, causal conclusions are not falsifiable. Instead, their validity is supported or refuted by indirect evidence of a logical or mechanistic nature. Despite this fundamental challenge, causal inference has seen an explosion in use over the past two decades, thanks in large part to the clarification of theoretical underpinnings, extensive testing and simulation, and the proliferation of powerful analytical tools. A search of https://app.dimensions.ai for the term “*causal inference*” showed that ∼190,000 out of 200,000 all-time hits (∼93%) were published after the year 1999 (4). Thus, it is clear that causal inference techniques are an exceptionally useful and important modern analytical tool.

### 1.3 Causal Identification

We will not recapitulate the entire field of causal inference here, since there are countless outstanding resources written by experts in the field. For a relatively non-technical yet rigorous and highly entertaining overview, we direct the reader to The Book of Why (Pearl and Mackenzie, 2018). The process of causal identification starts with proposing an underlying assumed model. One way such a model can be described is in terms of a directed acyclic graph (DAG). A DAG consists of a set of nodes, representing variables, connected to one or more other nodes by arrows. These arrows represent direct causes of one variable on another. The simple diagram A → B means: “*A causes B*” (e.g., the wind causes the leaves to flutter, not the other way around, as the 6-year-old version of the first author believed). Representing a set of causal hypotheses as a DAG has several strengths, including clear visual communication and the ability to rigorously define requirements for the estimation of causal effects. The latter is largely the result of the work of Pearl, and is the key tool in this study for examining causal effects related to walking impairments (5). A DAG also allows the identification of conditional independencies – partial correlations that must be zero (or small, in practice). These independencies can be tested to examine the plausibility of the proposed DAG. Satisfying the conditional independencies is a necessary, but not sufficient condition to show accuracy of the proposed model.

#### Total and Direct Effects

We will be examining both direct and total effects in this study. The *direct effect* of exposure A on outcome B is the effect passing through the causal arrow A → B. However, there may also be indirect effects of A on B. For example, perhaps the diagram consists of the paths A→B and A→M→B. In this case, A has a direct effect on B and an indirect effect on B, mediated by M. The *total effect* of A on B accounts for causal effects flowing through all paths – both direct and mediated.

#### Adjustment sets

Given a causal diagram we need a way to isolate the causal effect of a variable A (sometimes called an “*exposure*” or a “*treatment*”) on variable B (sometimes called an “*outcome*”). This is done by finding a set of variables called an adjustment set. The effect of A on B is causal after conditioning for the adjustment set, which consists of the other variables that influence the effect of A on B. In an experimental setting, this can be achieved by controlling for the adjustment set variables through experimental design. In an observational setting, we cannot control events. Instead, we include the adjustment set variables in a model. There are mathematically rigorous rules that allow us to find adjustment sets or determine that no such set exists (5). The adjustment set required to determine total effects may be different from the one required to determine direct effects. A proper adjustment set needs to be determined for each variable, in order to avoid misrepresenting a non-causal effect for a causal one – called the “*Table 2 Fallacy*” (6).

**Table 1.** Age, Sex, and Diagnosis

| Variable | N = 9,504 |
| --- | --- |
| Age, Median (IQR) | 10.5 (7.7 – 13.8) |
| Diagnosis, n (%) |  |
| CP | 6,667 (70) |
| Dev Variant | 597 (6.3) |
| Myelo | 314 (3.3) |
| TBI | 207 (2.2) |
| Other | 1,719 (18) |
| Sex, n (%) |  |
| Female | 4,247 (45) |
| Male | 5,257 (55) |

**Table 2.** History of Prior Treatement

| Variable | N = 9,504 |
| --- | --- |
| Prior SDR, n (%) | 2,242 (24) |
| Prior Neurolysis, n (%) | 3,850 (41) |
| Prior FDO, n (%) | 2,359 (25) |
| Prior TDO, n (%) | 1,220 (13) |
| Prior Foot Bone, n (%) | 1,716 (18) |
| Prior DFEO, n (%) | 162 (1.7) |
| Prior Adductor, n (%) | 891 (9.4) |
| Prior Foot Soft Tiss, n (%) | 1,309 (14) |
| Prior Calf, n (%) | 2,446 (26) |
| Prior Psoas, n (%) | 782 (8.2) |
| Prior Hamstrings, n (%) | 1,164 (12) |
| Prior Patellar Adv, n (%) | 342 (3.6) |
| Prior Rectus Transfer, n (%) | 814 (8.6) |

### 1.4 Causal Estimation

We use a computational model to turn the causal arrows in the DAG into numerical values. To do this we first represent the variables, which are abstract concepts like mobility, in the form of numbers (e.g., 1.35, 2.7, 42, …) or labels (e.g., mild, moderate, severe, …). The concept of the variable and its realization as a number or label will be considered exchangeable. If there is a well-understood mechanism underlying the causal path, the computational model can be explicit. For example, suppose a causal diagram has a direct arrow from A → B, and we know the effect of A on B is linear. Then we can model the path as *B* = *β* ∗ *A*. In this case, the causal effect is simply the coefficient *β*. If only the world was so simple.

The relationships we are examining in the model we are proposing do not generally have well understood mechanistic relationships that can be expressed in algebraic formulae. Also, the number of relationships as well as their complexity and unknown multi-way interactions presents a challenge. So, rather than proposing explicit relationships for each causal path, we take the approach of using a flexible computational engine that can accurately model these relationships.

We choose to use Bayesian Additive Regression Trees (BART) as the computational engine (7). Using BART for causal inference has been shown to produce accurate and precise estimates of causal effects across a wide range of real and synthetic datasets (8, 9). The reason for this exceptional performance derives from BART’s ability to flexibly recreate the shape of the outcome’s response surface.

#### Interpreting Results

In a linear regression it is straightforward to interpret an effect – it is the coefficient of the variable in the regression equation (*β* in the above example). A disadvantage of BART is that, as a “*black box*” machine learning algorithm, extracting interpretable causal effect magnitudes requires additional analysis. There are many proposed solutions to this problem, including partial dependence plots and accumulated local effects. In this study we have chosen to use Shapley values to represent the causal effect magnitudes (10). A variable’s Shapley value is the amount that it contributes to the model prediction for the outcome. Shapley values are frequently computed relative to the sample mean for the outcome – in which case the sum of the sample mean outcome plus all Shapley values for all variables equals the predicted outcome. Shapley values are not necessarily constant, but instead can depend on other covariates. As an analogy, we return to the linear regression example above A → B where *B* = *β* ∗ *A*. In this example *β* is the causal effect and has a constant value. But what if the value of *β* depended on the value of *A*? In that case we would have *B* = *β*(*A*) ∗ *A*. In fact, in a more complex situation, *β* could depend on several variables, not just *A*. In what follows, we present Shapley values three ways. If *S*_*age*_ is the Shapley value for the variable age, we can compute and display the mean absolute Shapley value over all observations (importance plot). We can also examine how age affects the Shapley value by displaying *S*_*age*_(*age*) (dependence plot). Lastly, we can compute Shapley values for important variables on an individual basis (case studies).

In the following sections we will introduce a model for understanding the causes and consequences of walking impairments. We will describe the model in detail and present five examples where the model is implemented using data collected during routine clinical gait analysis assessments.

## 2 Materials and Methods

### 2.1 The Model

The proposed causal model consists of 10 nodes and 23 primary causal paths [**Figure 1**]. For clarity, the depiction of the model omits some details that are described below and may be found in an included *Rmarkdown* file (n.b., we will use an italicized, monospaced font with a grey background to indicate software). The model captures causal relationships at a snapshot in time. In other words, all nodes, including *History* (n.b., nodes will be capitalized and displayed in italics throughout), are evaluated at *time* = *t*. For example, a patient may present with a hamstrings contracture and walk in a crouch gait pattern. The model allows us to examine the effect of hamstrings contracture on crouch gait. However, it is possible that continuing to walk in such a pattern could result in worsening of hamstring contracture, or perhaps the patient might undergo a hamstring lengthening surgery to improve the contracture. These changes would be reflected in a model of *time* = *t* + 1.

**Figure 1.**
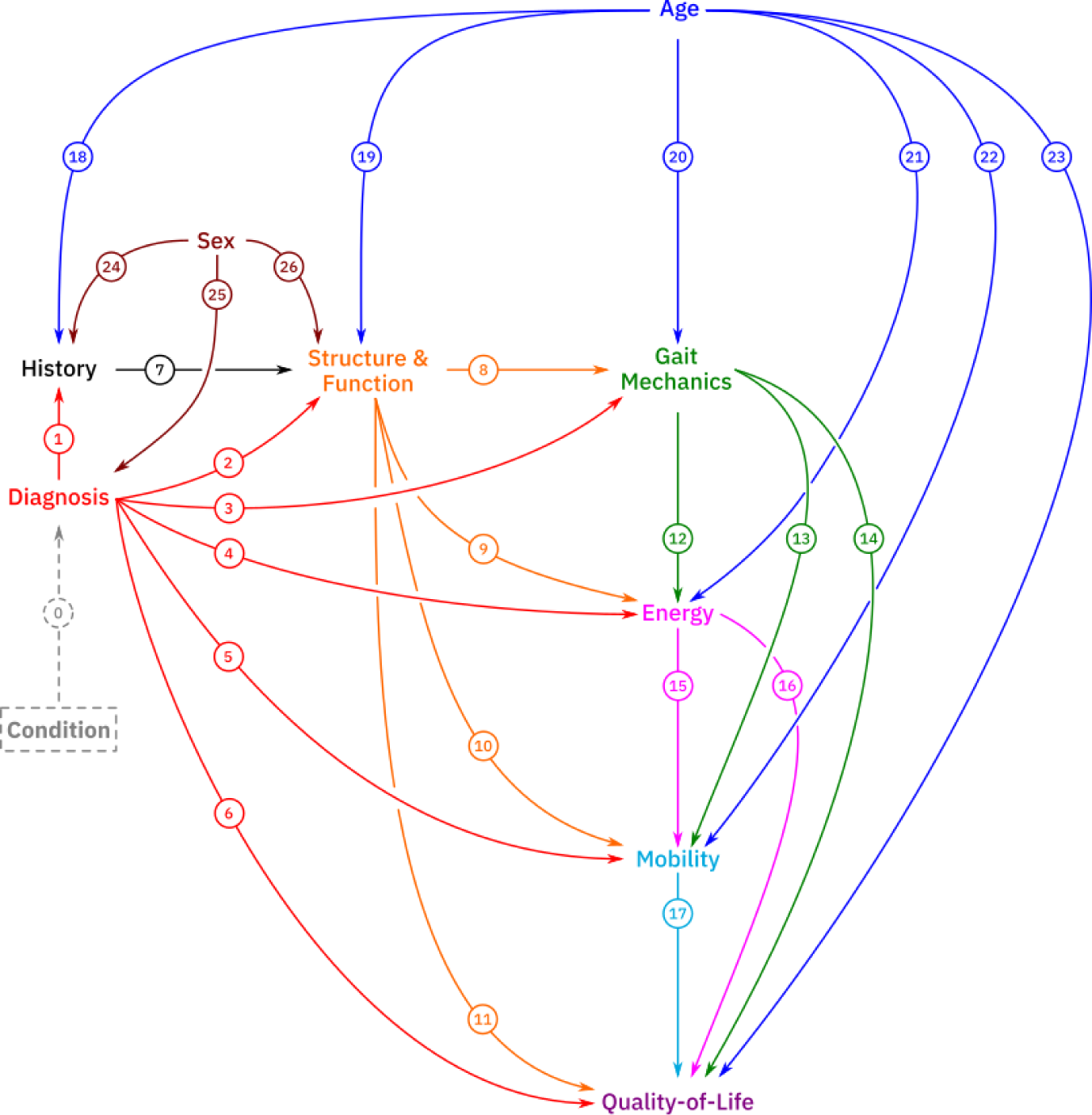
The model is presented here in schematic form. The proposed model nodes include the (latent) *Condition*, *History*, *Sex*, *Structure and Function*, *Mobility*, *Energy*, and *Quality-of-Life*. Extensive details of proposed nodes are found in the text, and paths in Appendix 1. A full implementation of the model is included as an *Rmarkdown* file model along with a de-identified dataset.

### 2.2 Model Nodes

*Condition* is a latent variable capturing the exact details of the patient’s medical condition. Note that *Condition* is a placeholder term for anything causing a change in body structure or function. This could include injuries, diseases, genetic mutations, malnutrition, aging, etc.

*Diagnosis* is a surrogate for the *Condition* variable. We do not expect *Diagnosis* alone to capture the full details of the patient’s condition. However, this coarse parameter allows us to adjust for some of the missing condition-dependent variables we do not measure.

*History* includes events that can significantly influence body structures and function – such as prior treatments. Since the model uses the exam date as a starting point, we do not model the causes of the prior treatments. Therefore, within the *History* node, we connect prior treatments to a latent variable of unknown common causes (*Prior*_*j*_←*Unknown*→*Prior*_*k*_, for all *j,k*). Note that this detail is not shown on the schematic version of the causal model, but can be found in the included *Rmarkdown* file.

*Structure and Function* is shorthand for Body Structures and Functions as described in The International Classification of Functioning, Disability and Health (ICF) framework (11). We are specifically thinking of variables like spasticity (ICF category b750 – Motor reflex functions and b735 – Muscle tone functions), motor control (b760 – Control of voluntary movement functions), strength (b730 – Muscle power), range-of-motion, bony alignment, muscle contracture, and joint health (s740 – Structure of pelvic region, s750 – Structure of lower extremity, s760 – Structure of the trunk, and s770 – Additional musculoskeletal structures related to movement). The *Structure and Function* node is the core of the proposed causal model since elements of this node are the direct targets of treatments, and treatment effects are largest at this level (12).

*Gait Mechanics* are the elements of dynamics pertaining to the motion of segments and joints during gait (usually walking, but running, skipping, and other gaits could be considered). These data can come from three-dimensional gait analysis, video, or observational analysis, depending on the questions being investigated. *Gait Mechanics*, as we mean it, appears in the ICF as both a body function (b770 – Gait pattern functions) and as an activity (d450 – Walking). When choosing variables to represent *Gait Mechanics*, additional choices need to be made about the relationships among those variables. Variables in this node are related to one another through definition as well as the geometric and physical constraints of the lower extremity and walking. Given the topology of the assumed causal model, it can be shown that these relationships do not impact important total causal effect sizes (e.g., *Structure and Function* → *Gait Mechanics* or *Gait Mechanics* → *Energy*).

However, they do impact implied conditional independencies. For example, if minimum hip flexion and hip flexion range of motion during gait (related algebraically) are included as part of the Gait Mechanics node, then a shared cause must be added (i.e., minimum hip flexion ← Unknown → range-of-motion hip flexion). We therefore add an unknown factor to this node that is connected to all *Gait Mechanics* variables (*Gaint Mechanics*_*j*_←*Unknown*→*Gaint Mechanics*_*k*_ for all *j,k*). Note that this detail is not shown on the schematic version of the causal model (**Figure 1**) but can be found in the included *Rmarkdown* file.

Cadence, step-length, and speed do not appear explicitly in the causal model. Instead, they are considered part of *Gait Mechanics*, but merit special attention. Speed is the product of cadence and step-length. These are part of ICF Walking (d450 – Walking), which is defined as “*Moving along a surface on foot, step by step, so that one foot is always on the ground, such as when strolling, sauntering, walking forwards, backwards, or sideways*.” Alert readers will note that the ICF defines walking, in part, as “*walking*”. The trio of cadence, step-length, and speed are not independent. Any two can be chosen freely and fully determine the third. We choose cadence and step-length as the causes of speed since, to walk (1) we turn muscles on and off in a sequence and intensity that generates joint movements, (2) the joint movements result in a step of a given length, and (3) we repeat the step-length generating activation pattern at a given cadence. Thus, we choose to treat cadence and step-length as the independent causal variables. Including speed is redundant.

However, there are meaningful effects that are more sensibly expressed as speed effects rather than as combined cadence and step-length effects. For example, the inability to keep up with peers while walking in the community, a frequent patient complaint, is most clearly expressed as a speed problem. Other effects are more naturally expressed in terms of cadence and step-length, such as the effect these variables have on energy (13). Thus we include cadence, step-length, and speed in the causal model and include the condition that cadence → speed ← step-length. It is important to keep this redundancy in mind when working with the model.

*Energy* is the metabolic energy required for walking (ICF b5409 – General metabolic functions unspecified or b789 – Movement functions other specified and unspecified). We recommend against using energy per unit distance – often called “*energy cost*”, which is obtained by dividing metabolic power by walking speed. Problems arise at slow speeds, where energy cost is dominated by the speed^-1^ term, and the impact of the metabolic power term becomes trivial.

*Mobility* is a broad term referring to a range of functions and abilities related to moving around within the home or community (ICF d410-429 – Changing and maintaining body position, d430-449 – Carrying, moving, and handling objects, d450-469 – Walking and moving, and d47-489 – Moving around using transportation). The elements of mobility that are important are patient-specific, and influenced by culture, environment, and many other factors.

*Quality-of-Life* may be clear conceptually but is a vexingly difficult quantity to define and measure. For one patient, walking faster may have a large impact on their quality-of-life. For another, walking speed may not matter, but gait appearance does. Despite the central importance of this measure, it is the most likely to be influenced by factors not represented in the diagram, such as social relations, socio-economic status, psychiatric disturbances, job status, and so on. There are some successful and widely used measures for this domain, such as the World Health Organization Quality of Life Assessment and the Diener Satisfaction with Life Scale (14, 15). We leave *Quality-of-Life* in the diagram as a reminder of its central importance, while simultaneously acknowledging the limitations of the model regarding this node.

### 2.3 Causal Paths

Each path in the model indicates the possible existence of a causal effect. It is only necessary to find one possible example of the path for it to be included in the model. The magnitude of the effect may turn out to be trivially small, but that is for the data to show. Given this situation, one might ask “*why not allow possible causal paths between all pairs of variables?*”. The answer is that such a fully connected model produces no implied conditional independencies, and therefore, cannot be falsified. It also ignores the nature of the true causal relationships among variables, which may result in the introduction of nonexistent and biasing paths. We do not indicate if an effect is linear, quadratic, or some other form. Nor do we describe the existence or nature of interactions. Those determinations are part of the causal estimation described in section 1.4, Causal Estimation. Brief examples for each of the 23 causal paths in the proposed model can be found in Appendix 1.

### 2.4 Missing Nodes and Paths

No causal model can include every node and path. Some missing nodes that seem important include socio-economic status, cognition, family dynamics, internal motivation, pain, or other medical conditions. Missing nodes and paths have two possible impacts. First, they may increase unexplained variance in an outcome measure. Second, and generally more importantly, they may bias the causal estimates. It is difficult to predict whether the missing nodes and paths in a model are biasing or not. There are methods to estimate how much bias missing paths would need to introduce to change causal conclusions (16). We will not perform these sorts of analyses in this study.

### 2.5 Model Realization

#### Data

This study was reviewed and authorized by the University of Minnesota institutional review board review (STUDY00012420). All experiments were performed in accordance with relevant guidelines and regulations. Informed consent for use of medical records was obtained at the time of service from all participants or their legal guardians. An option to rescind this permission is offered to patients at every visit to our center.

We used data extracted from our laboratory’s historical clinical database collected between 1994 and 2022. Data consisted of measures collected for clinical standard-of-care evaluations.

For the computational models, missing continuous data was left as missing. Missing categorical data was neither omitted nor imputed, but instead assigned its own category (“*Missing*”). For causal effects dependency plots, we did not plot the “*Missing*” category, since we are not modeling the pattern of missingness and hence this category has no clear causal meaning. Handling of missing data in future applications of the model (e.g., hypothesis testing) must be carefully considered.

#### Variables

To turn the conceptual causal model into a practical tool, we needed to realize the nodes as measured values. The variables contained in the model nodes were chosen to be comprehensive and commonly measured in clinical gait analysis centers to promote replication efforts and model extensions.

*Diagnosis*: Assigned diagnosis lumped into the categories cerebral palsy, myelomeningocele, developmental variants, traumatic brain injury, and other.

*History*: Prior treatments lumped into the categories selective dorsal rhizotomy, neurolysis (e.g., botulinum toxin type A or phenol injection), rectus femoris transfer, femoral derotation osteotomy, tibial derotation osteotomy, distal femoral extension osteotomy, foot and ankle bone surgery, psoas lengthening, hamstrings lengthening, adductor lengthening, calf muscle lengthening, patellar advancement, foot and ankle soft-tissue surgery. These 13 account for around 90% of the treatments performed on patients in our clinical database. The causal effects of these treatments have been evaluated previously (18).

*Sex*: Sex as indicated by the patient’s medical record.

*Age*: Age ex-vivo. Note that when dealing with very young individuals or incipient walkers, it might be more appropriate to consider an adjusted age that accounts for premature birth.

*Structure and Function*:

- <u>Maximum range-of-motion</u>: hip flexion, hip extension, hip abduction (knee extended), popliteal angle (unilateral), knee flexion, knee extension, extensor lag, ankle dorsiflexion (knee extended), ankle plantarflexion (measured with handheld goniometry).
- <u>Long bone torsion</u>: trochanteric prominence angle test, bimalleolar axis angle (measured with manual goniometry).
- <u>Weightbearing foot alignment</u>: forefoot ab/adduction, forefoot in/eversion, hindfoot var/valgus, midfoot position (subjectively assessed)
- <u>Spasticity</u>: overall lower extremity spasticity defined as first component score from polychoric principal component analysis applied to spasticity of the adductors, hamstrings, hip flexors, plantarflexors, posterior tibialis, and rectus femoris (19). The resulting measure is a Z-score with respect to the sample.
- <u>Strength</u>: overall lower extremity strength defined as first component score from polychoric principal component analysis applied to manual muscle strength testing of abdominal, back extensors, hip abductors, hip adductors, hip extensors, hip flexors, knee extensors, knee flexors, ankle plantarflexors, anterior tibialis, posterior tibialis (20). The resulting measure is a Z-score with respect to the sample.
- <u>Static selective motor control</u>: overall lower extremity static selective motor control defined as first component score from polychoric principal component analysis applied to subjectively assessed measures of isolated static motor control (0-fully patterned, 1-partially patterned, 2-normal) of abdominal, back extensors, hip abductors, hip adductors, hip extensors, hip flexors, knee extensors, knee flexors, ankle plantarflexors, anterior tibialis, posterior tibialis. The resulting measure is a Z-score with respect to the sample.
- <u>Dynamic motor control</u>: The Dynamic Motor Control Index during Walking (21). This measure requires electromyography data. For many years we did not routinely collect electromyography data during post-treatment visits, resulting in a greater amount of missing data for this measure.

#### Gait Mechanics

Kinematic parameters derived from three-dimensional gait analysis: mean pelvic tilt, pelvic tilt range-of-motion (ROM), minimum stance-phase hip flexion, mean stance-phase hip rotation, minimum stance phase knee flexion, knee flexion at initial contact, swing phase knee flexion ROM, maximum swing phase knee flexion, mean stance knee rotation , ankle dorsiflexion at initial contact, mean stance ankle dorsiflexion, mean swing ankle dorsiflexion, mean stance foot progression, dimensionless step-length, dimensionless cadence, dimensionless speed. All kinematics are obtained from the Vicon plug-in gait model with functional hip and knee axis (22, 23). For data collected prior to 2006, we did not compute a functional knee axis and therefore do not report a knee rotation value. Speed, cadence, and step-length nondimensionalized following Hof (24).

#### Energy

Net dimensionless metabolic walking power z-score with respect to speed-matched typically-developing controls (25). For simplicity we will refer to this as metabolic walking power.

#### Mobility

Functional Assessment Questionnaire Transform (FAQt) (26). Demonstrated mobility measures like the GMFM-66 would be preferable, but these data are not available for most of the patients in this study. The FAQt has been shown to be strongly correlated with the GMFM-66 (r ∼ .60).

#### Quality-of-Life

Domain scores from the GOAL survey: Activities of Daily Living and Independence, Body Image and Self-Esteem (27).

#### Software

We use the *dagitty* package in *R* to build the DAG (17). Programming details are in the included *Rmarkdown* file (electronic addendum).

#### Deriving Adjustment Sets

We derived adjustment sets using the provided functions in *dagitty*.

#### Computational Engine

We use BART models to compute the causal effects. For outcome variables expressed as continuous or dichotomous variables we use the *bartMachine* package (28). Note that outcomes expressed as polytomous variables can be predicted using the *BART* package (Sparapani, Spanbauer, and McCulloch 2021). We use default settings for all BART models, which have been shown to be nearly optimal across a wide range of conditions. We found only ∼1 - 3% improvement in out-of-sample performance from cross-validation optimized models.

#### Quantifying Causal Effect Magnitude with Shapley Values

Shapley values were computed using the *fastShap* package (29). We present Shapley values in three different forms: (1) importance plots display mean absolute Shapley values and give an overall picture of the causal importance of a variable, (2) dependence plots show how the Shapley value varies over the domain of the causal variable and (3) case studies show Shapley values of chosen variables for individual patients.

### 2.6 Plausibility of the Model

Model plausibility was tested using implied conditional independencies. In the case of the proposed model, most of these involved large numbers of conditioning variables (2 tests with 0 conditioning variables, 48 with 1, 165 with 15, 280 with 20, and 1 with 38). The calculations are unreliable in this situation, so we only evaluated conditions with 0 or 1 conditioning variables (30).

Furthermore, many of the tests did not provide meaningful plausibility information since they were essentially tests of the relationship between speed, cadence, and step length, which is fulfilled by definition. To compute test values we first generated a polychoric correlation matrix for the variables using the *lavaan* package (31). Note that this step was necessary since the implied conditional independencies involved a mix of continuous and ordered data. The diagnosis variable was further lumped into two categories – CP (∼80% of observations) and Other. This dichotomization was necessary since diagnosis is an unordered category.

### 2.7 Examples

We demonstrate the utility of the proposed causal model with five examples. The purpose of these examples is to show how to use the model, highlight the model’s potential clinical and scientific value, and provide some face validity. There are many different combinations of exposures and outcomes we could consider – each producing dozens of graphs and tables. Rather than giving an encyclopedic treatment here we are providing an *Rmarkdown* file and a demonstration data set of 300 deidentified observations so that readers can run the *Rmarkdown* file to create facsimiles of the plots presented here and can modify the code to explore, enhance, test, or utilize the causal model. For each example we (1) chose an exposure node and outcome measure, (2) determined the proper adjustment set for valid causal inference, (3) built a BART model for the outcome based on the adjustment set, and (4) examined a combination of importance plots, dependence plots, and case studies.

#### Example 1: Total effects of *Structure and Function* on mean stance foot progression

The effect of impairments at the *Structure and Function* level on impairments at the *Gait Mechanics* level is the primary problem of interest in most clinical gait studies and directly drives treatment decisions. The broader goals of treatment may include improving *Energy*, *Mobility*, or *Quality-of-Life*, but the treatment assignment process is largely based on assumptions about how structural impairments cause gait impairments. These assumptions are *ad hoc*, rarely based on evidence, and essentially never based on causal evidence, since the gait literature contains (almost) none. To demonstrate the application of the proposed model to this causal path we examined the total effects of *Structure and Function* on in- and out-toeing, which is a common problem of concern to patients and is frequently treated with surgery. We quantified in- and out-toeing by mean stance foot progression. The adjustment set consists of variables belonging to the *Structure and Function*, *Age*, and *Diagnosis* nodes [Appendix 2].

#### Example 2: Total effects of *Structure and Function* on Functional Assessment Questionnaire Transform (FAQt)

*Mobility* is important to a patient and is often discussed in relation to treatment decisions. We have chosen to quantify *Mobility* using the FAQt. Note that some components of the FAQt are clearly mediated by *Gait Mechanics* (e.g., overall walking ability and climbing stairs) and some are not (e.g., kicking, hopping, and jumping). The adjustment set consists of variables belonging to *Structure and Function*, *Age*, and *Diagnosis* nodes. Note that it would be wrong to adjust for *Gait Mechanics*, which is a mediator (i.e., there is a path consisting of *Structure and Function* → *Gait Mechanics* → *Mobility*) [Appendix 2].

#### Example 3: Total effect of *Gait Mechanics* on metabolic walking power

The power demand for walking in children with cerebral palsy is high – averaging over twice that of typically developing controls. Common treatments show limited effectiveness to reduce power demands. In a previous study, overall kinematic deviations were shown to contribute substantially to elevated power demands (32). We quantify *Energy* as net (walking – resting) dimensionless power expressed as a z-score with respect to speed matched controls (metabolic walking power) (25). By examining the effects of *Gait Mechanics* on metabolic walking power we can determine which specific kinematic deviations are the most impactful. The adjustment set consists of variables belonging to the *Gait Mechanics*, *Structure and Function*, *Age*, and *Diagnosis* nodes [Appendix 2].

#### Example 4: Direct effect of *Age* on metabolic walking power

A particular challenge with observational data is disentangling direct effects of age from effects due to other factors. including treatment. In this example, we examine the direct effect of age on metabolic walking power. By examining the direct effect of age on metabolic walking power we can quantify the effect that maturation has on metabolic walking power, independent of associated changes in *Structure & Function*, *History*, or *Gait Mechanics*. Unlike the previous examples, where we estimated total effects, here we examine the direct effect of *Age*. This is an effect that is impossible to observe – hence an ideal situation for causal inference methods. The direct effect isolates the developmental role of *Age* from the concomitant impact it has on factors like gait mechanics and treatment decisions, which result in indirect causal effects. The adjustment set consists of variables belonging to the *Diagnosis*, *Structure and Function*, and *Gait Mechanics* nodes [Appendix 2]. For this example, we analyzed only individuals diagnosed with cerebral palsy.

#### Example 5: Total effects of (1) *Structure and Function* on Activities of Daily Living and Independence and (2) *Gait Mechanics* on Body Image and Self-Esteem

Independence and self-esteem are important aspects of *Quality-of-Life*. The GOAL survey provides validated self-report measures from these domains. We use the GOAL’s Activities of Daily Living and Independence domain score as a measure of independence, and the Body Image and Self-Esteem score as a measure of self-esteem. We estimate the total effects of *Structure and Function* on the Activities of Daily Living and Independence score and *Gait Mechanics* on the Body Image and Self Esteem score. The adjustment set for the total effects of *Structure and Function* on Activities of Daily Living and Independence consists of variables belonging to the *Age*, *Diagnosis*, and *Structure and Function* nodes. The adjustment set for the total effect of *Gait Mechanics* on Body Image and Self-Esteem consists of variables belonging to the *Age*, *Diagnosis*, *Structure and Function*, and *Gait Mechanics* nodes [Appendix 2].

For examples 1 through 3 we computed Shapley values for a random sample of 1500 observations, which is around 1/6^th^ of the data and gives a representative picture of the response without making the plots overly dense. For example 4 we *also* included all observations younger than 4 years and older than 15 years in order to have sufficient density across ages. For example 5 we selected all available data.

## 3 Results

### 3.1 Data

The database query resulted in 9504 individuals (Tables 1-5). We selected the affected side (unilateral diagnosis) or the left side (bilateral diagnosis) from each individual for further analysis. We have not seen a meaningful impact from the use of bilateral data in this model or previous causal treatment effect models, though there may be situations where using bilateral data makes sense.

**Table 3.**
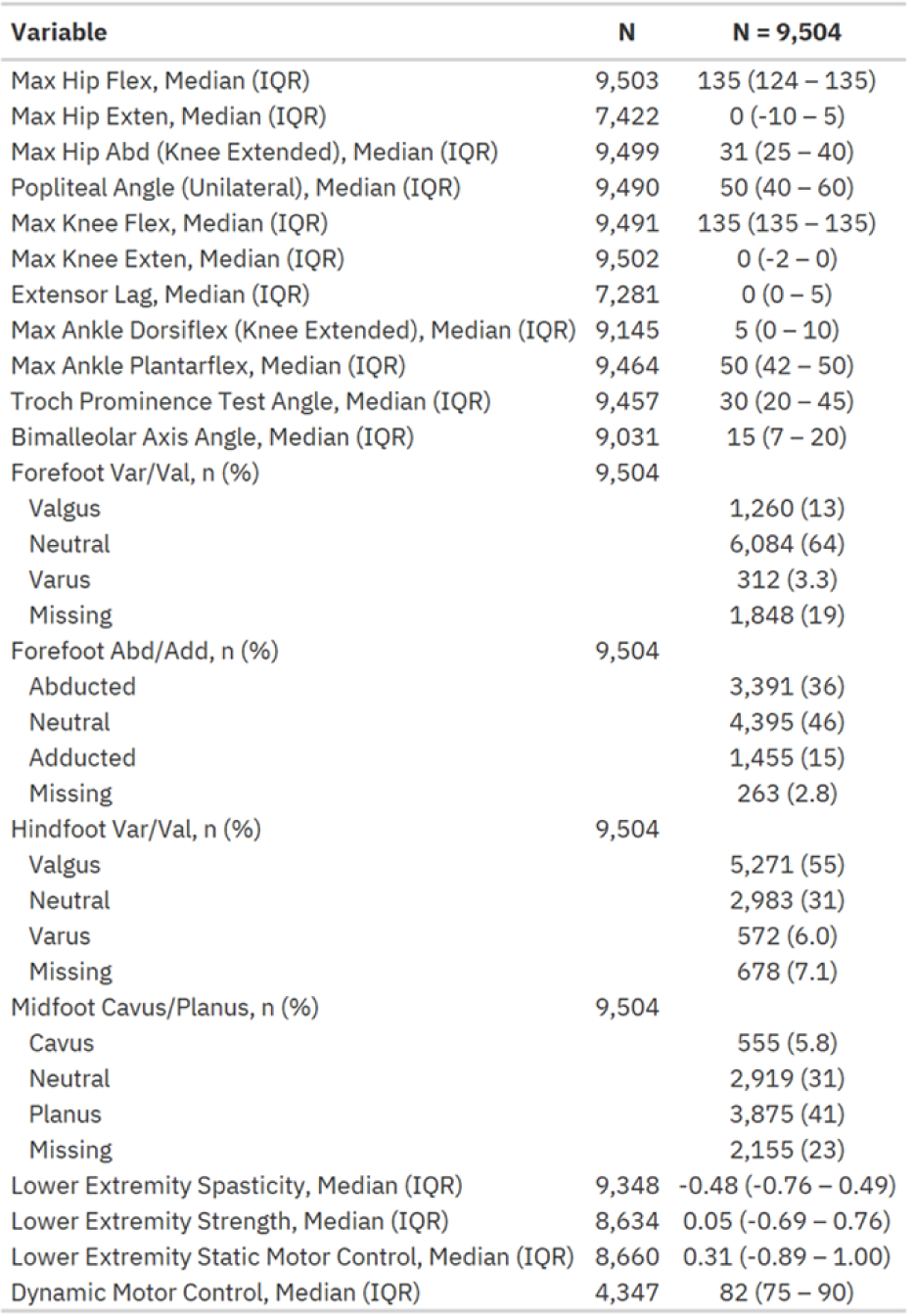
Structure and Function

**Table 4.**
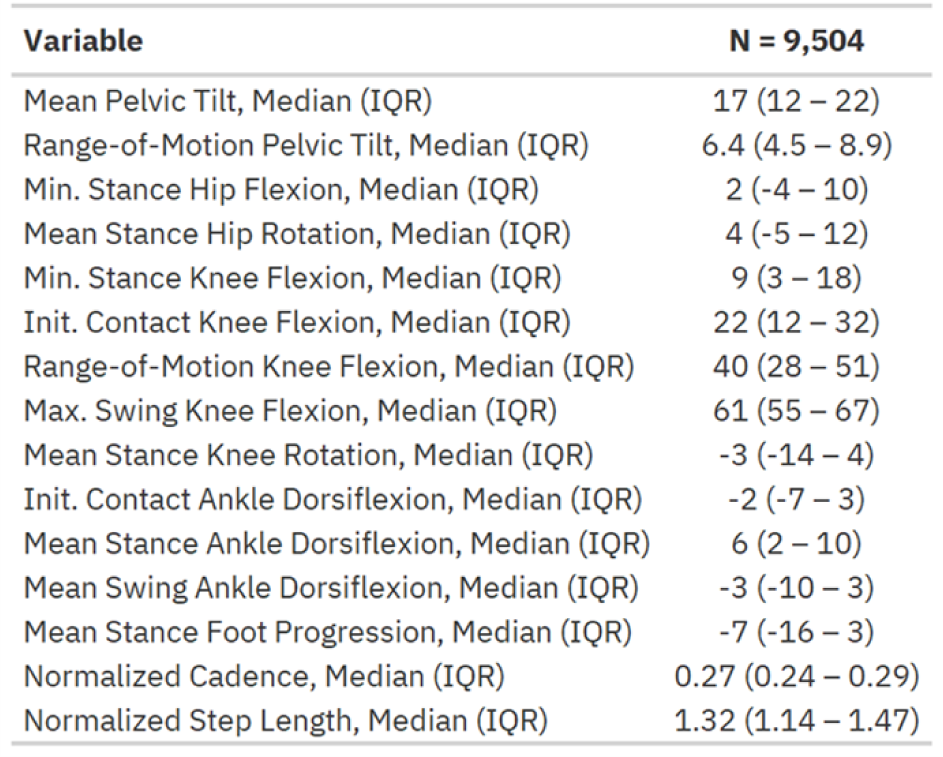
Gait Machanics

| Variable | N = 9,504 |
| --- | --- |
| Mean Pelvic Tilt, Median (IQR) | 17 (12 – 22) |
| Range-of-Motion Pelvic Tilt, Median (IQR) | 6.4 (4.5 – 8.9) |
| Min. Stance Hip Flexion, Median (IQR) | 2 (-4 – 10) |
| Mean Stance Hip Rotation, Median (IQR) | 4 (-5 – 12) |
| Min. Stance Knee Flexion, Median (IQR) | 9 (3 – 18) |
| Init. Contact Knee Flexion, Median (IQR) | 22 (12 – 32) |
| Range-of-Motion Knee Flexion, Median (IQR) | 40 (28 – 51) |
| Max. Swing Knee Flexion, Median (IQR) | 61 (55 – 67) |
| Mean Stance Knee Rotation, Median (IQR) | -3 (-14 – 4) |
| Init. Contact Ankle Dorsiflexion, Median (IQR) | -2 (-7 – 3) |
| Mean Stance Ankle Dorsiflexion, Median (IQR) | 6 (2 – 10) |
| Mean Swing Ankle Dorsiflexion, Median (IQR) | -3 (-10 – 3) |
| Mean Stance Foot Progression, Median (IQR) | -7 (-16 – 3) |
| Normalized Cadence, Median (IQR) | 0.27 (0.24 – 0.29) |
| Normalized Step Length, Median (IQR) | 1.32 (1.14 – 1.47) |

**Table 5.**
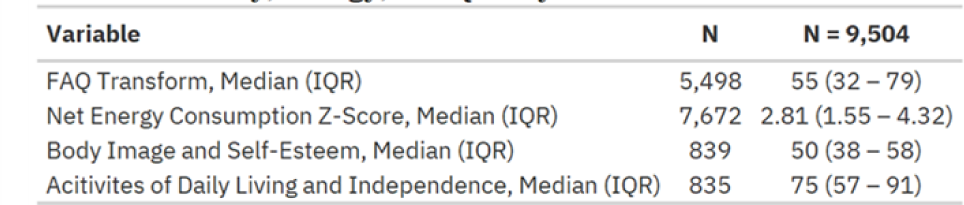
Mobilty, Energy, and Quality-of-Life

| Variable | N | N = 9,504 |
| --- | --- | --- |
| FAQ Transform, Median (IQR) | 5,498 | 55 (32 – 79) |
| Net Energy Consumption Z-Score, Median (IQR) | 7,672 | 2.81 (1.55 – 4.32) |
| Body Image and Self-Esteem, Median (IQR) | 839 | 50 (38 – 58) |
| Activities of Daily Living and Independence, Median (IQR) | 835 | 75 (57 – 91) |

### 3.2 Implied Conditional Independencies

As noted in the methods, due to the number of variables in the model, many implied conditional independencies could not be tested. The 50 conditions that could be tested exhibited partial correlations less than 0.20.

### 3.3 Examples

For each example, observations where the outcome was missing were excluded resulting in a reduced sample size in examples 1-5 (N = 9504, 5498, 7672, 7672, and 835/839, respectively).

#### Example 1: Total effects of *Structure and Function* on mean stance foot progression

As expected, tibial torsion (estimated by bimalleolar axis angle) and femoral anteversion (estimated by trochanteric prominence test angle) were the two largest causes of mean stance foot progression [**Figure 2**]. The next three largest causes of meaningful magnitude were related to foot deformity.

**Figure 2.**
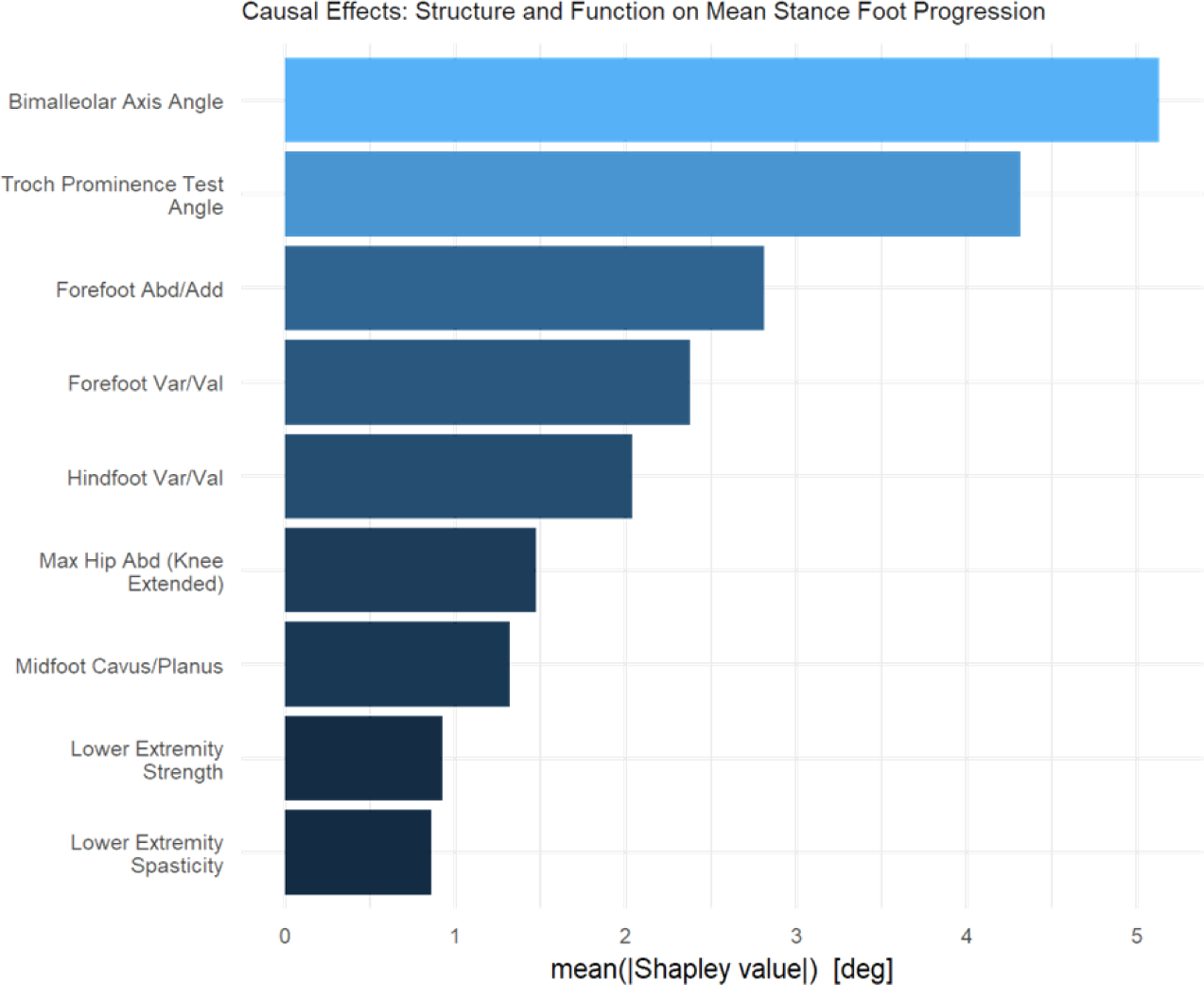
The nine most important causal effects of *Structure and Function* on mean stance foot progression. Unsurprisingly, torsions of the tibia and femur are the two largest contributors. Foot deformities comprised the next three largest causes.

The dependence of mean stance foot progression on long bone torsion appears to be approximately linear, with slopes of around 0.6 and 0.3 degrees of mean stance foot progression per degree of torsion for the tibia and femur, respectively [**Figure 3**]. We also see the expected relationship at the forefoot, with an abducted forefoot causing external foot progression and an adducted forefoot causing internal foot progression. Identifying these foot-related effects is reassuring since they are sensible, but also pleasantly surprising, since these measures tend to be quite coarse and noisy.

**Figure 3.**
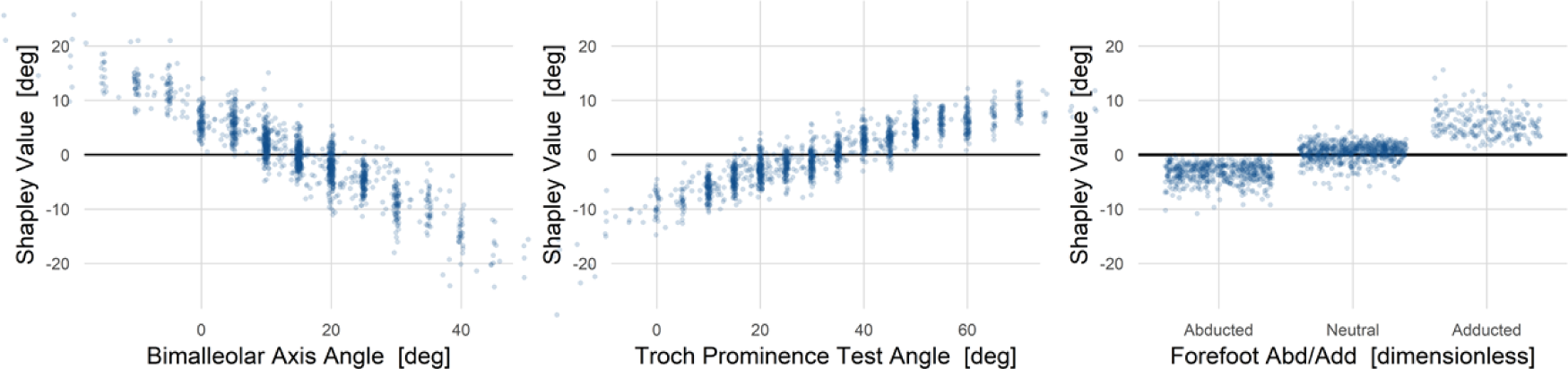
The three *Structure and Function* variables with largest causal effect on mean stance foot progression. Long bone torsions (femur, tibia) and forefoot deformity contribute meaningfully. Note that for many years measurements were only recorded at five-degree increments, leading to vertical striations in the left and middle panels.

As an aside, recall that Shapley values are expressed with respect to the mean over the sample, so dependence plots cross the zero line at the sample mean of the corresponding exposure value. In other words, visual inspection of **Figure 3** shows that the sample mean bimalleolar axis angle is ∼15°, trochanteric prominence test angle is ∼35°, midfoot cavus/planus is neutral, and forefoot abduction/adduction is neutral.

Four examples from individuals, two with significant in-toeing (top row) and two with significant out-toeing (bottom row), show different *Structure and Function* profiles leading to similar in- and out-toeing [**Figure 4**]. This has obvious clinical importance since different treatments would be considered each of these cases.

**Figure 4.**
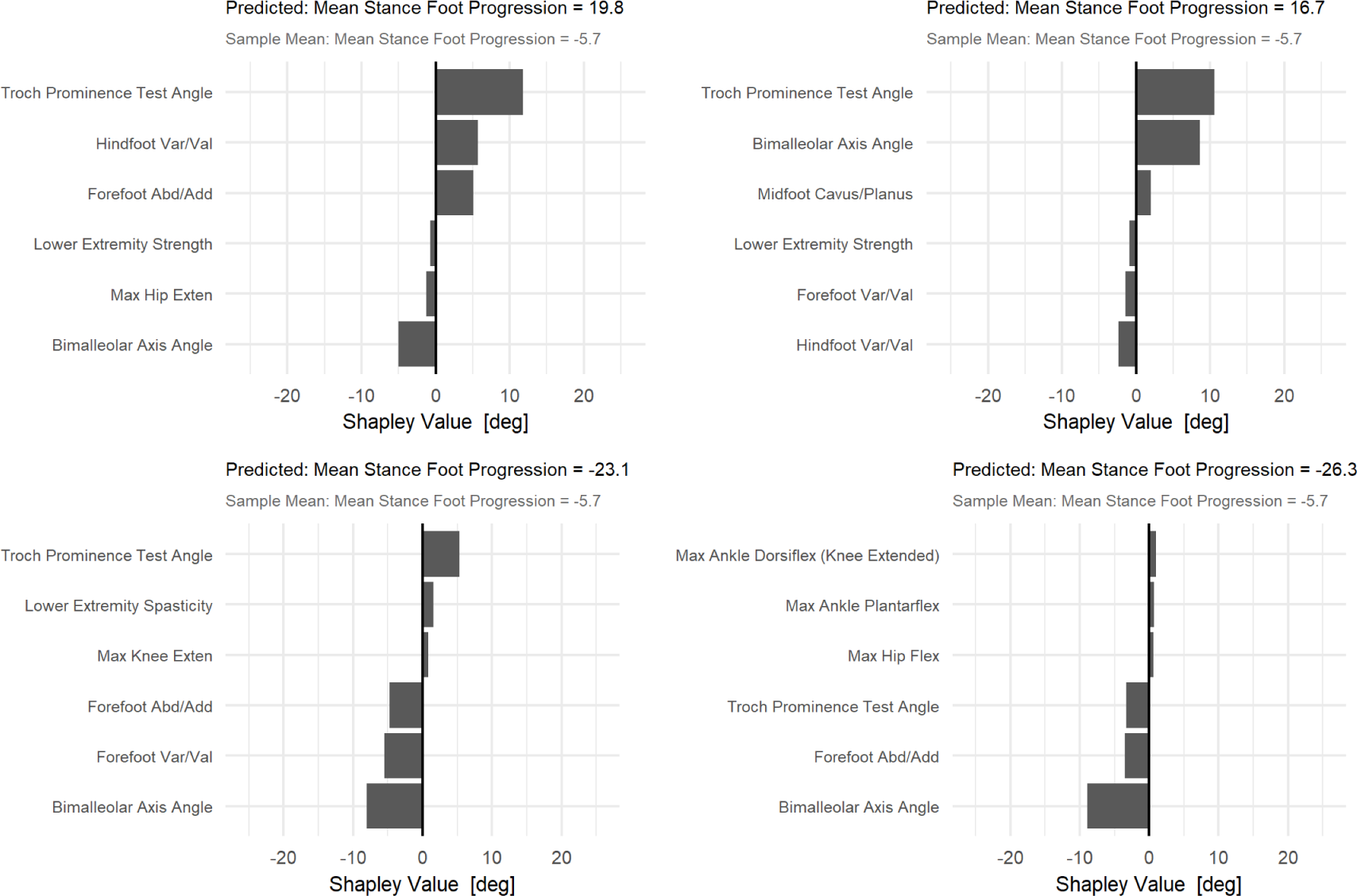
Case studies showing the three largest positive (in-) and negative (out-toeing) causal factors. The top row shows patients with significant in-toeing. On the top-left, the primary cause is excess femoral anteversion, with additional contributions from hindfoot varus and forefoot adduction. On the top-right femoral anteversion and tibial torsion both contribute. The bottom row shows patients with significant out-toeing. On the bottom left, external tibial torsion and forefoot deformity cause the out-toeing. On the bottom right, both femoral retroversion and external tibial torsion contribute meaningfully, along with forefoot abduction.

#### Example 2: Total effects of *Structure and Function* on Functional Assessment Questionnaire Transform (FAQt)

Strength and motor control (static and dynamic) were the most important causal contributors to FAQt [**Figure 5**].

**Figure 5.**
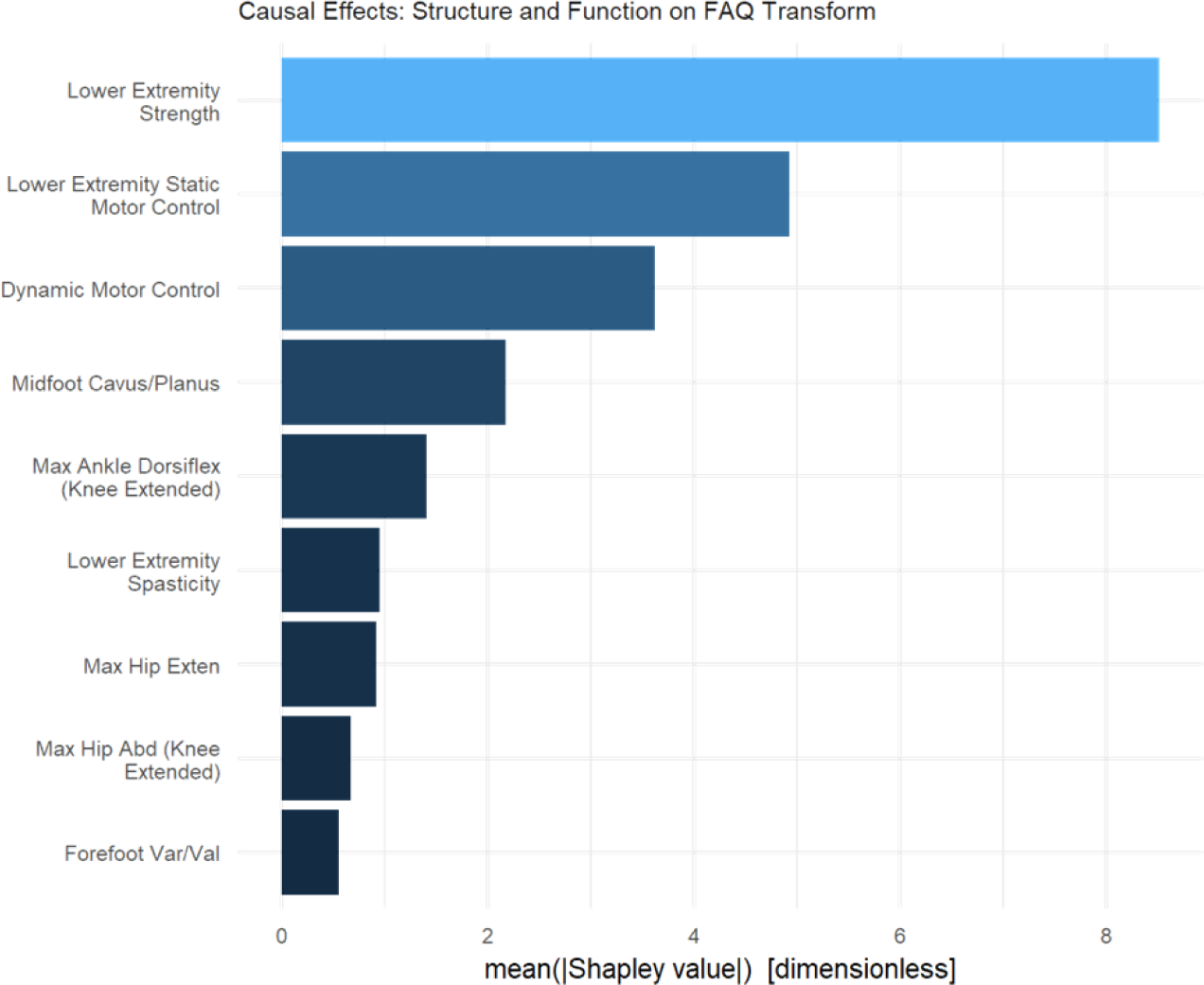
The effect of *Structure and Function* variables on mobility as measured by the FAQt. Strength and motor control (static and dynamic) are the main contributors.

The dependence plots suggest that the difference between poor overall strength (-1 which is 1 SD weaker than sample average) and good strength (+1which is 1 SD stronger than sample average) is 20 points, or 2 SD on the FAQt scale [**Figure 6**]. While this is a large effect, the likelihood of improving overall strength by 2 SD through any intervention is low. The motor control effects are nearly as large but improving motor control is also an extremely challenging therapeutic goal.

**Figure 6.**
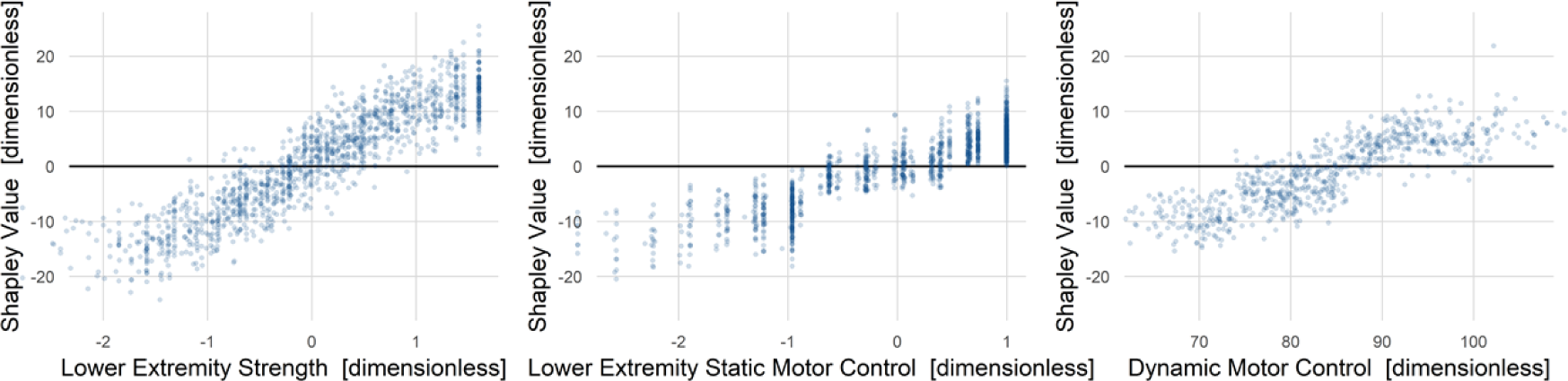
The effect of *Structure and Function* variables on mobility as measured by the FAQt. Strength and motor control (static and dynamic) constitute the top three contributors.

All highly mobile patients resemble one another. They are strong and have good motor control. Those with poor mobility also fit a consistent prototype of poor strength and poor motor control. Individual examples of both high and low FAQt scores typify this finding [**Figure 7**].

**Figure 7.**
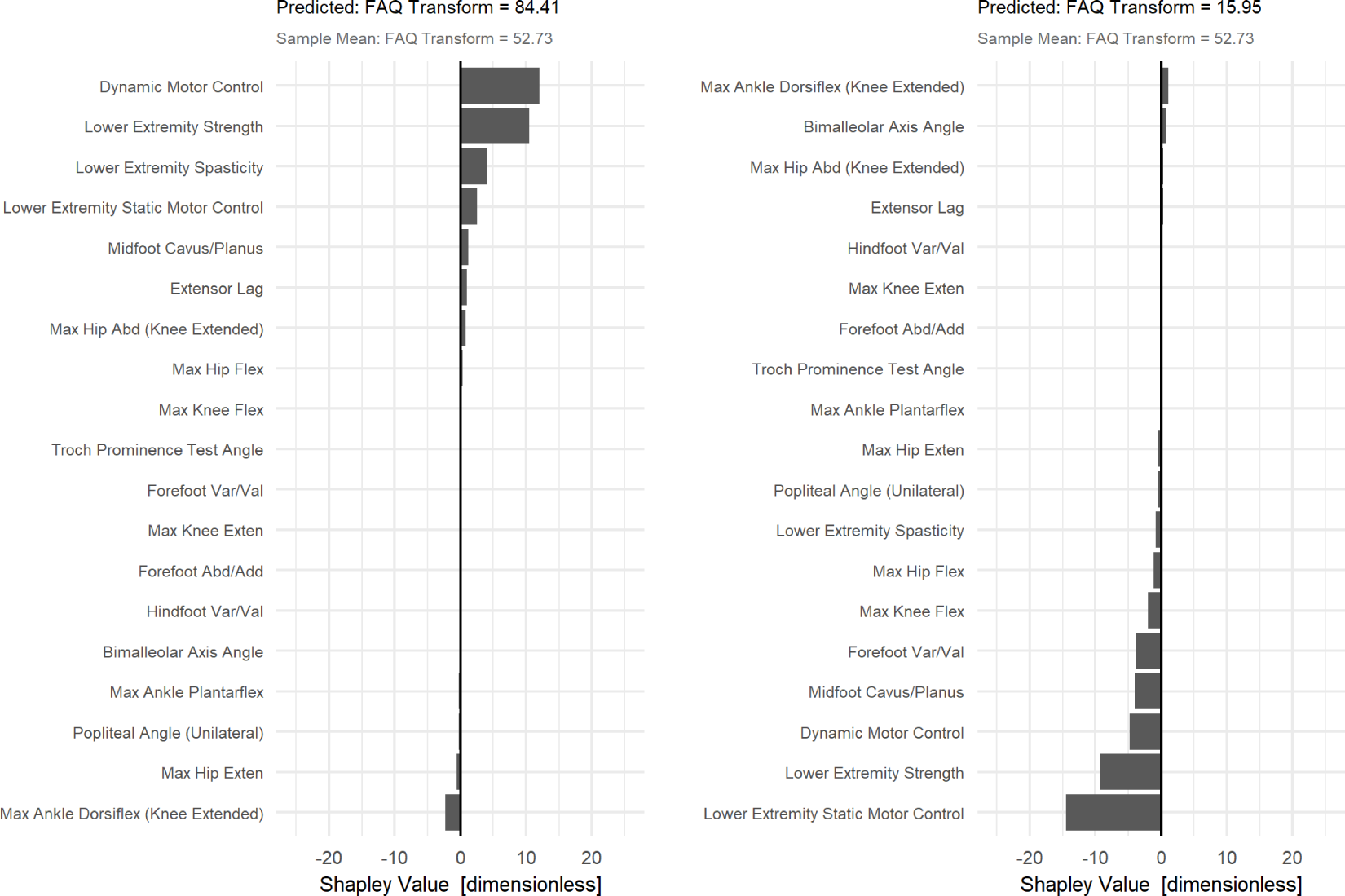
Two case studies showing the 10 largest positive and negative causal factors of FAQt. High mobility is caused by good strength and motor control, low mobility is caused by the opposite.

#### Example 3: Total effect of *Gait Mechanics* on metabolic walking power

The largest causal contributor to metabolic walking power was initial contact knee flexion, which was twice as impactful as the next closest cause [**Figure 8**]. Elements of stance phase knee flexion and ankle dorsiflexion were the next most important causes – equal in magnitude to one another.

**Figure 8.**
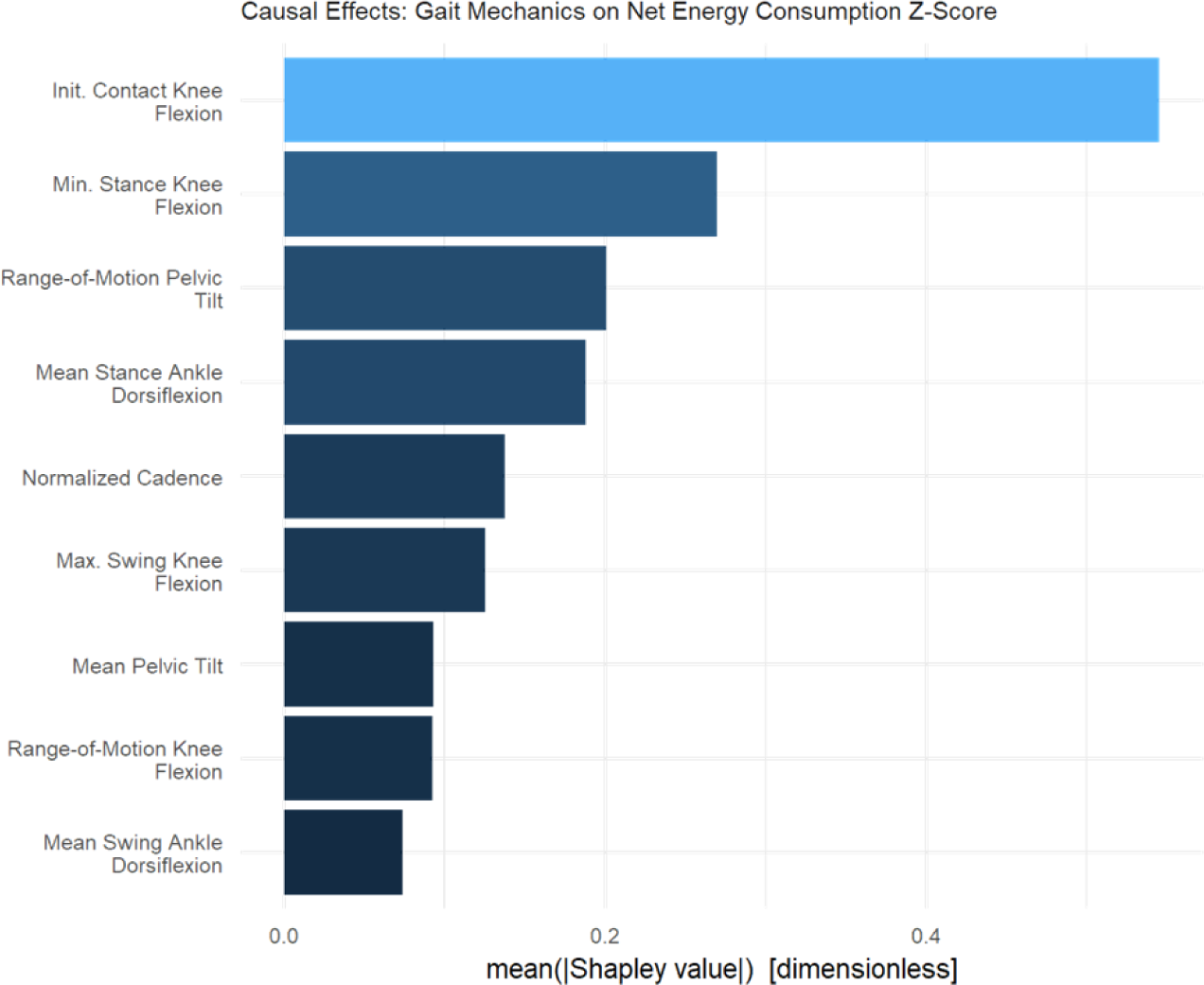
Knee flexion at initial contact has a causal impact twice as large as the next largest contributors – stance phase features of the knee and ankle.

Shapley value dependence plots showed that initiating ground contact with the knee in a flexed position has a substantial metabolic walking power penalty – around 1.5 SD difference between contacting with 10° vs. 50° of flexion [**Figure 9**].

**Figure 9.**
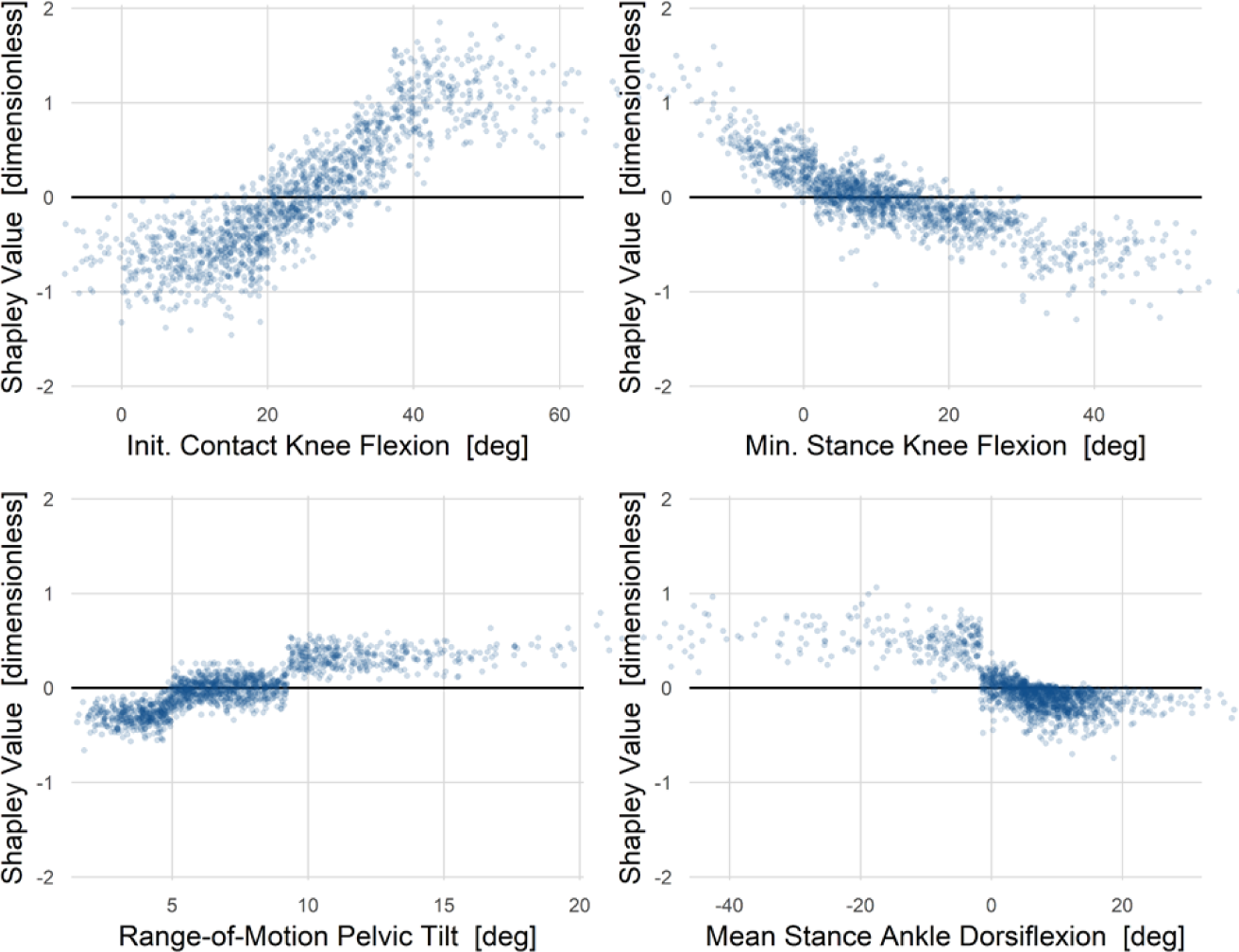
The effect of *Gait Mechanics* on walking power. Being flexed at contact is imparts a substantial energetic penalty. Knee hyperextension is energetically costly, but maintaining a flexed position is not. This somewhat counterintuitive finding is discussed further in text. A large pelvis range-of-motion and stance phase ankle plantarflexion were also substantial causes of high walking power – though much smaller than the effects of knee kinematics.

It is interesting to note that walking in crouch (minimum stance knee flexion > 25°) seems to provide an energetic benefit. Previous studies, both experimental and analytical, have reported mixed and inconclusive findings regarding the relationship between energy and crouch. It is possible that this is because they failed to control or adjust for the proper confounders (33, 34). Walking in a plantarflexed position also contributes to elevated metabolic walking power. The apparent threshold effect for plantarflexion may be a consequence of using a tree-based computational model, or it may reflect a true mechanism related to the way that a heel strike promotes an energetically efficient step-to-step transition (13). The disruption of passive walking dynamics may also be the mechanism underlying the power penalty caused by knee hyperextension.

After observing the minimum stance-phase knee flexion result, we hypothesized that this reflected the energetic penalty for extending a loaded and flexed knee and thus raising the body center-of-mass (recall that the initial contact position is accounted for in the model). Such a pattern is sometimes called “*jump gait*”. We explored this hypothesis by stratifying the minimum stance phase knee flexion plot by initial contact knee flexion, binned into mild, moderate, and severe categories [**Figure 10**]. We see that, in support of our hypothesis, only individuals who land in severe crouch derive a metabolic walking power benefit from remaining in crouch. Not all these individuals derive the metabolic walking power benefit, indicating that additional causal factors play a role. The results of the model are suggestive of a mechanism but need to be tested experimentally or through simulations.

**Figure 10.**
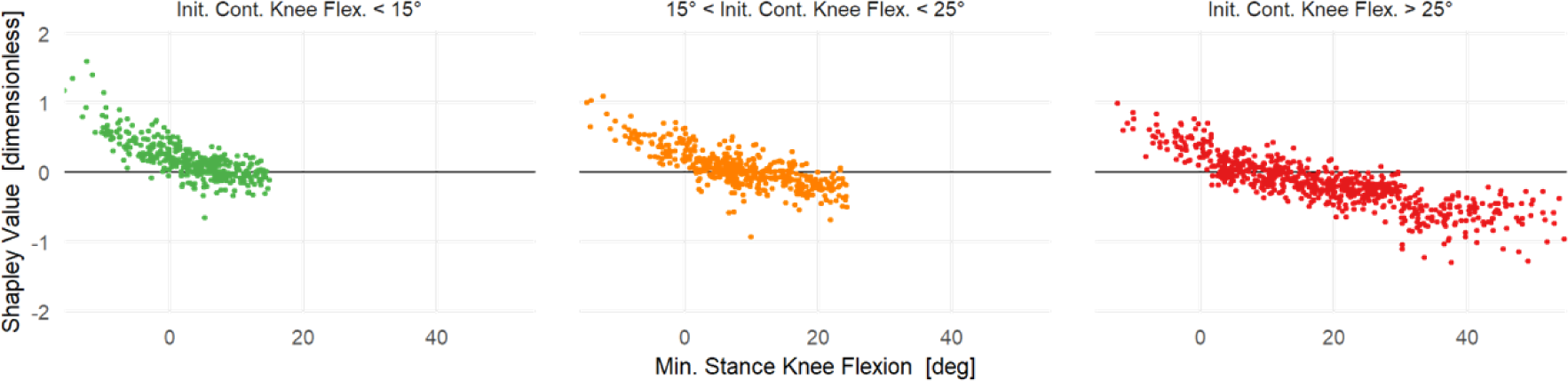
The Shapley value for minimum stance knee flexion on walking power, stratified by initial contact knee flexion. The results support the hypothesis that *remaining* in crouch is energetically beneficial, and thus only individuals who land in a severely crouched position exhibit a negative Shapley value (lower walking power). Not all these individuals derive the energy benefit, suggesting a role for additional causal factors.

Two examples, each from individuals with high energy levels, show that different Gait Mechanics profiles can lead to similar levels of walking energy [**Figure 11**]. The two cases have identical dimensionless metabolic walking power, but one is in severe crouch while the other is in equinus and knee hyperextension.

**Figure 11.**
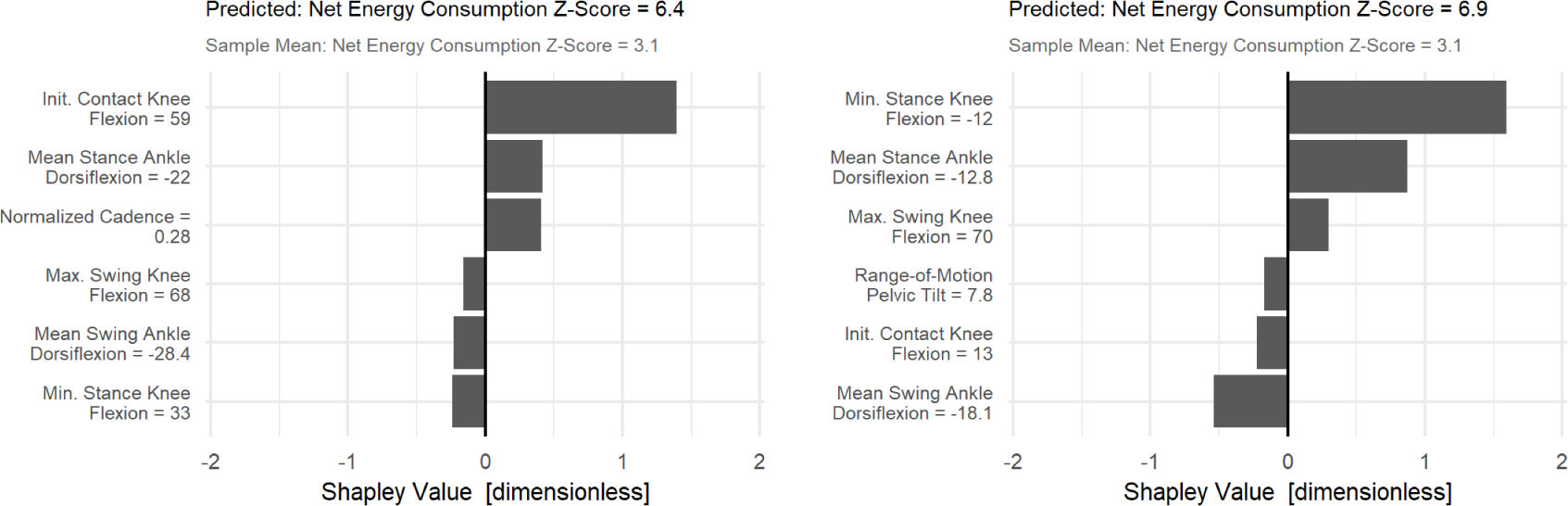
Two case studies in walking power showing the three largest positive and negative causal factors (the measured values of the causal factors have been added to the y-axis labels). The two patients depicted have energy between 6 and 7 SD above speed-matched typically developing controls. For the patient on the left initial contact knee flexion (59°) is an important cause of high energy. Minimum stance knee flexion for this patient (33°) is severe crouch – yet lowers the power demand. The hypothesized mechanism for this is described in the text. For the patient on the right, knee hyperextension and excessive ankle plantarflexion both contribute meaningfully to the elevated walking power.

#### Example 4: Direct effect of *Age* on metabolic walking power

Metabolic walking power undergoes a substantial reduction with age (∼ 1 SD) [**Figure 12**]. For clarity we have fit a generalized growth (Richards’) curve to the data (35). This results in an initial mean metabolic walking power of +0.49, final mean metabolic walking power of -0.42, midpoint of maturation of 10.1 years, and time constant of 0.49. The age dependence shows significant energy decrease (0.91 SD) occurring primarily between ages 5 – 12 years.

**Figure 12.**
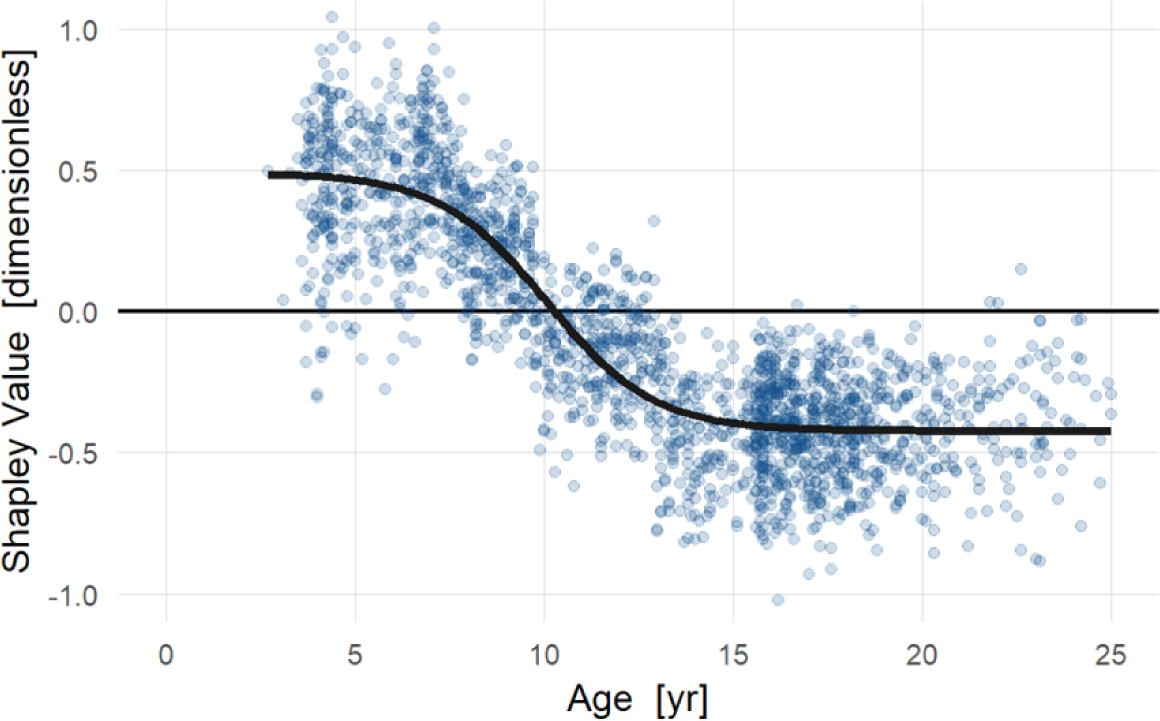
The direct effect of age on walking power shows meaningful maturation effects, accounting around 1 SD of walking power reduction. The period of rapid reduction occurs from around 7.5 – 12.5 years. Evaluation of treatment effects during this epoch needs to account for the direct effects of age.

An implication of this effect’s importance can be seen in outcomes of selective dorsal rhizotomy. Elevated metabolic walking power is commonly observed in children with cerebral palsy.

Historically, selective dorsal rhizotomy was believed to be effective at reducing metabolic walking power. Recent studies have shown that this is not true (36, 37). At our center the mean baseline and follow-up ages for selective dorsal rhizotomy evaluations are 6.3 and 7.8 years, which is during the period where age alone – independent of treatment – causes rapid reductions in metabolic walking power. Thus, the misattribution of age effects as treatment effects probably contributed to the erroneous ideas about selective dorsal rhizotomy and metabolic walking power.

#### Example 5: Total effects of *Structure and Function* on Activities of Daily Living and Independence *and Gait Mechanics* on Body Image and Self-Esteem

Strength and motor control (static and dynamic) are the largest *Structure and Function* variables affecting Activities of Daily Living and Independence [**Figure 13**]. As has been discussed earlier, these neurological factors are difficult to change with currently available treatments.

**Figure 13.**
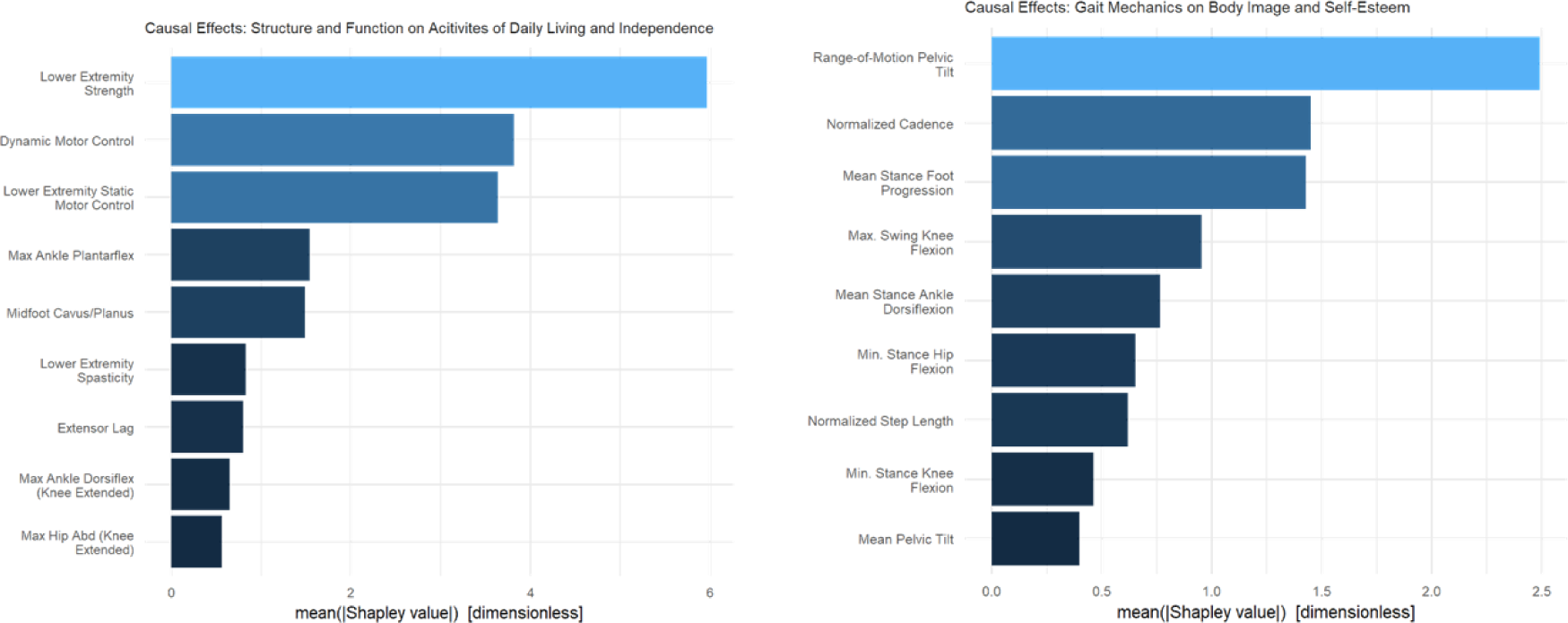
Average causal effects of *Structure and Function* on Activities of Daily Living and Independence (left) and *Gait Mechanics* on Body Image and Self-Esteem (right). Some of the important causal variables can be addressed with well- established treatments. For example, ankle plantarflexion (Activities of Daily Living and Independence) or mean stance foot progression (Body Image and Self-Esteem). Strength and motor control are more difficult to change using currently available treatments.

The next largest causal factors were related to ankle and foot deformity. Among *Gait Mechanics* variables affecting Body Image and Self-Esteem, large pelvic tilt range-of-motion, rapid cadence, and foot progression were the three largest causal factors. The model was able to identify detrimental effects on Body Image and Self-Esteem of both out- and in-toeing (external and internal foot progression) – with internal having a substantially larger impact [**Figure 14**]. Foot progression is effectively treated with femoral and tibial derotation osteotomies and bony foot surgery.

**Figure 14.**
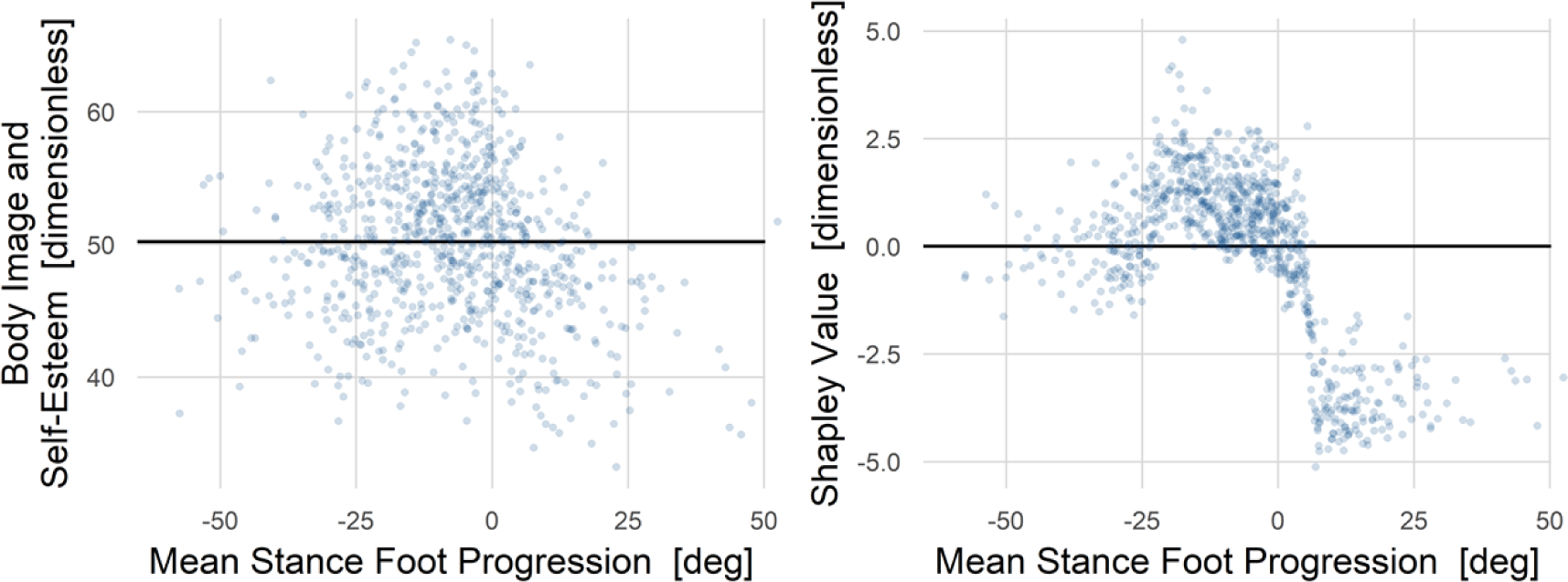
Raw (left) and causal (right) impact of in- and out-toeing on Body Image and Self-Esteem. The raw data is widely dispersed but is suggestive of detrimental effects when deviating from typical foot alignment (around -10°). These effects are much clearer in the causal dependence plot. Deviations in either direction from typical result in lower Body Image and Self-Esteem, but the impact of in-toeing (positive) is about twice as large as that of out-toeing. There is a precipitous drop starting around 0°, which is probably where in-toeing becomes easily discernable.

## 4 Discussion

### 4.1 Summary

The purpose of this study was to propose a comprehensive model for evaluating causal effects of walking impairments and their consequences. We have described the model, demonstrated its plausibility using data from a pediatric clinical motion analysis laboratory, and presented several ways that it can be used to test and expand our understanding of walking impairments. The model provides results and data that can further our understanding of the relationships between treatments and outcomes by identifying the important sources of an impairment. The model’s output reinforces some beliefs, provides useful details regarding others, and generates new hypotheses. The model can also be used to estimate causal effects for individuals, with obvious implications for supporting treatment decisions.

### 4.2 Examples

This is not a “*validation*” study, *per se*, and the examples were not meant to prove the model is correct. Nevertheless, it is important that the model produces results that provide an element of face validity by reproducing – perhaps with more detail and rigor – existing knowledge. The large impact of femoral anteversion and tibial torsion on in- and out-toeing is not surprising. However, in addition to confirming this well-known effect, the model estimates the functional form, showing that tibial torsion is about twice as impactful as femoral anteversion on a degree-for-degree basis. In- and out-toeing were also found to be detrimental to Body Image and Self-Esteem, a widely believed concept, but one that is not easily seen in raw data. In addition to clearly identifying this causal link, the model showed that in-toeing was around twice as impactful as out-toeing. Like the torsion findings, the direct effect of age on metabolic walking power is not novel, but the model demonstrates this effect using substantially different methods than have been used in the past.

Furthermore, we derive an explicit functional form for the age-dependence that can be used for adjusting observed treatment effects. The evaluation of metabolic walking power also confirmed the critical role of knee flexion. However, rather than finding a monotonic crouch → power relationship, we saw a subtle yet sensible interaction effect between initial contact knee position, maximum knee extension, and power. While the purported mechanism needs to be tested, uncovering unknown or poorly understood phenomena like this exemplifies an important role that causal models can play. We found that strength and motor control are the key causal factors for both *Mobility* and *Quality-of-Life* (Activities of Daily Living and Independence). This result agrees with clinical beliefs and suggests why correction of orthopedic impairments does not lead, on average, to improvements in mobility (18).

### 4.3 Interpreting Model Outputs

Shapley values are well-established, principled, and easily interpreted method for quantifying causal effects. Other commonly used methods are partial dependence plots, accumulated local effects plots, and individual conditional expectation plots. All these methods give more-or-less the same answer. This raises an important point regarding the sort of precision we expect from causal models. Many are familiar with the saying “*measure with a micrometer and cut with a hack-saw*”.

Here, we do not have that problem since we are essentially using a hacksaw for both measuring and cutting. In other words, our goal is not to identify effects down to the millimeter or degree, but to identify the main causal factors, rule out non-causes unjustified beliefs, and derive reasonable sign and magnitude estimates for causal effect sizes (38).

Shapley values estimate causal factors and therefore give useful guidance for treatment decisions. However, Shapley values do not necessarily predict changes associated with treatment. There are several reasons for this. Multiple variables may change with a treatment, leading to a different causal impact due to interactions. Furthermore, patients can compensate for changes introduced by treatment. For example, suppose a patient’s Shapley profile indicated in-toeing largely caused by femoral anteversion. Now suppose the patient underwent a femoral derotation osteotomy of magnitude matching the Shapley value. Finally, suppose the patient “*compensated*” after surgery by internally rotating their pelvis by half the amount of the derotation. In this case, the treatment outcome would only be half of the original causal effect of anteversion, despite a full orthopedic correction. Additional work is needed to quantify the relationships between causal effects and treatment outcomes.

### 4.4 Limitations

All causal models are proposals that can never be fully confirmed or refuted, and thus need to be evaluated critically. The minimal level of model checking involves testing implied conditional independencies to establish model plausibility. Most implied conditional independencies could not be evaluated for the proposed model due to the large numbers of conditioning variables. Those that could be tested supported the plausibility of the model. We know that there are missing paths in the model. Some of these missing paths could represent important causes while others could introduce bias. However, not every missing path leads to bias. For this to be the case, missing variables would, at a minimum, need to be a common cause of the exposure and outcome of interest. For example, cognition would need to affect *Structure and Function* and *Gait Mechanics* for its absence to bias the findings regarding torsion and in-toeing. Furthermore, even if biasing paths exist, it is not clear how large an effect such a bias would have. This question can be studied with sensitivity analysis.

The *Diagnosis* node is crude. The most obvious next element to add to this node is a measure of severity. Those involved in CP research will immediately notice the absence of the Gross Motor Function Classification System (GMFCS), which is often viewed – explicitly and implicitly – as a severity measure. We have left the GMFCS out of the model for two main reasons. The first reason is that we do not have similar scales for other diagnoses and would thus introduce significant imbalance in missing values. This is a relatively minor problem since missing values can be handled seamlessly by *bartMachine*. The second reason, which is more important and more intriguing, is that it is unclear whether the GMFCS level is a cause or an effect. The original description suggests GMFCS as a cause – a fixed value that is a surrogate for severity (39, 40). However, in practice, a GMFCS level is assigned based on functional skills like climbing stairs or carrying objects. This creates a paradox, since the skills dictating the GMFCS assignment are caused by *Structure and Function* and *Gait Mechanics* impairments. In other words, it is not clear whether the causal arrow is GMFCS → *Structure and Function*/*Gait Mechanics* or *Structure and Function*/*Gait Mechanics* → GMFCS. Given the lack of formal causal modeling in CP research, it is not surprising that this paradox has not been discussed previously. The proper causal model would appear to at least contain GMFCS ← Severity → *Structure and Function*/*Gait Mechanics*, with Severity a latent variable. We have experimented with other severity measures such as age of walking onset, which seems to work well. However, establishing the validity of such a severity measure is beyond the scope of this study.

In this study we have only examined continuous outcomes. The basic methodology applies directly to categorical outcomes (binary, ordered, polytomous), but for ordered and polytomous outcomes it is a necessary and straightforward modification to use a different BART engine, since *bartMachine* does not handle ordered and polytomous outcomes. We have successfully used the *BART* package for these types of outcomes (41).

### 4.5 Future Directions

As stated at the outset, this model is a proposal that is meant to be critically evaluated, validated or refuted, altered, and improved over time. Such improvements might include the introduction of new nodes, variables, and paths. We are particularly interested in modeling the effects of social determinants of health, though we recognize the monumental challenge this presents. *Quality-of-Life* is affected by many factors not in the model, such as employment, social relations, and other health conditions, to name a few. Nevertheless, the results (sensibly) suggest that certain impairments of *Structure and Function* and *Gait Mechanics* reduce *Quality-of-Life*. The current model does not include pain. Pain is clearly an important phenomenon, but it is unclear how to properly include it. For example, pain can be a cause of *Gait Mechanics* (antalgic gait caused by osteoarthritic pain) or pain can be an effect of *Gait Mechanics* (e.g., knee pain caused by pre-patellar pressures present in crouch gait).

The replication crisis in science is real. There are vanishingly few examples of important clinical gait studies being replicated or refuted (42). To encourage open and transparent scientific practice, we are providing a detailed *Rmarkdown* file and a sample dataset. We expect that this will greatly ease the burden of implementing this model in other centers and will help researchers identify model shortcomings and propose possible improvements.

Causal inference is inherently ambiguous since we cannot observe multiple realizations of the same person with different characteristics. We have proposed a model, demonstrated its utility, and provided limited face validity. We have made our methods transparent and easy to implement and scrutinize by peers. It is important to design experiments and simulations that test the predictions and underlying assumptions of this model. That difficult task must be the source of future efforts.

## Data Availability

All relevant data are within the manuscript and its Supporting Information files.

---

**Appendix 1.**
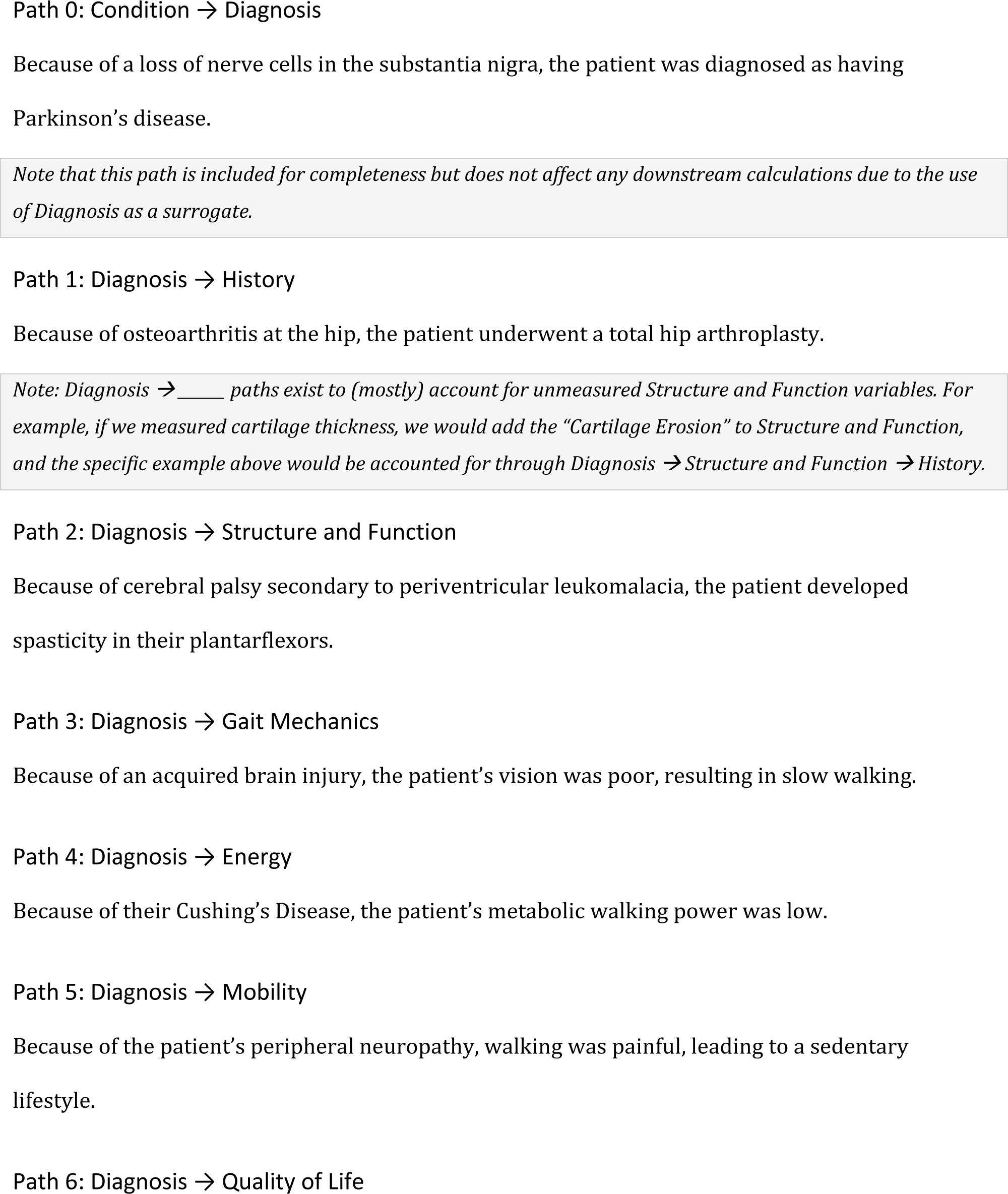
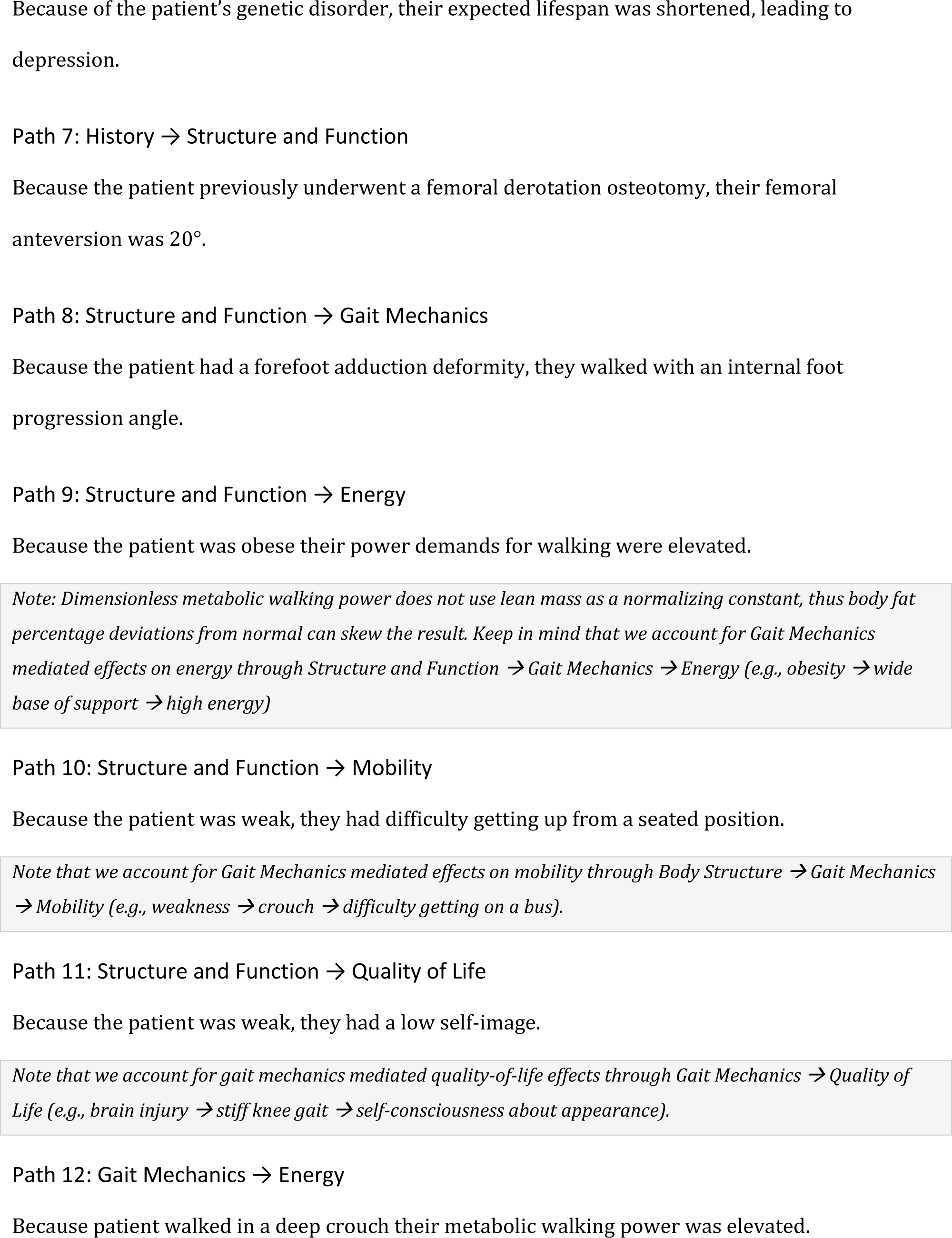
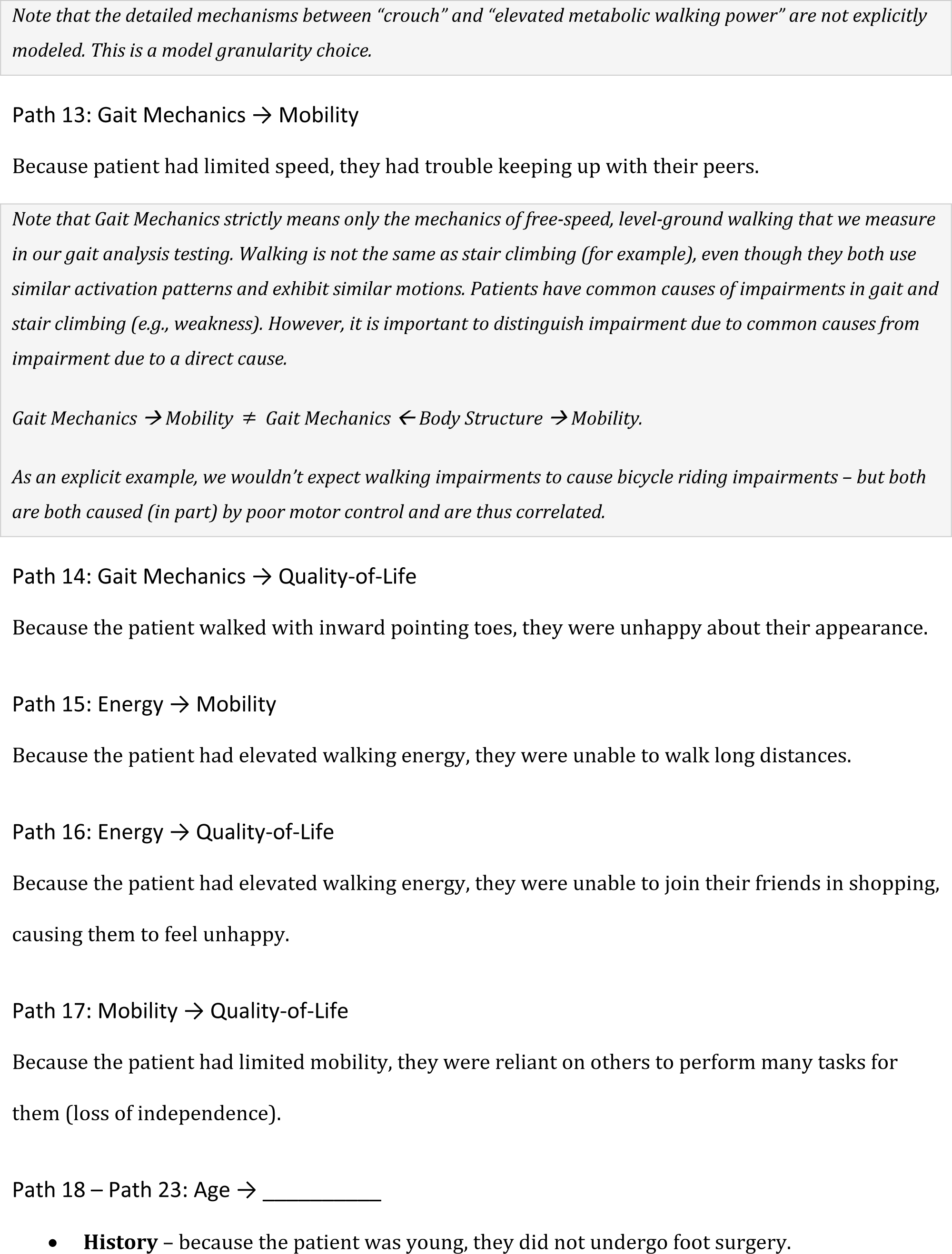
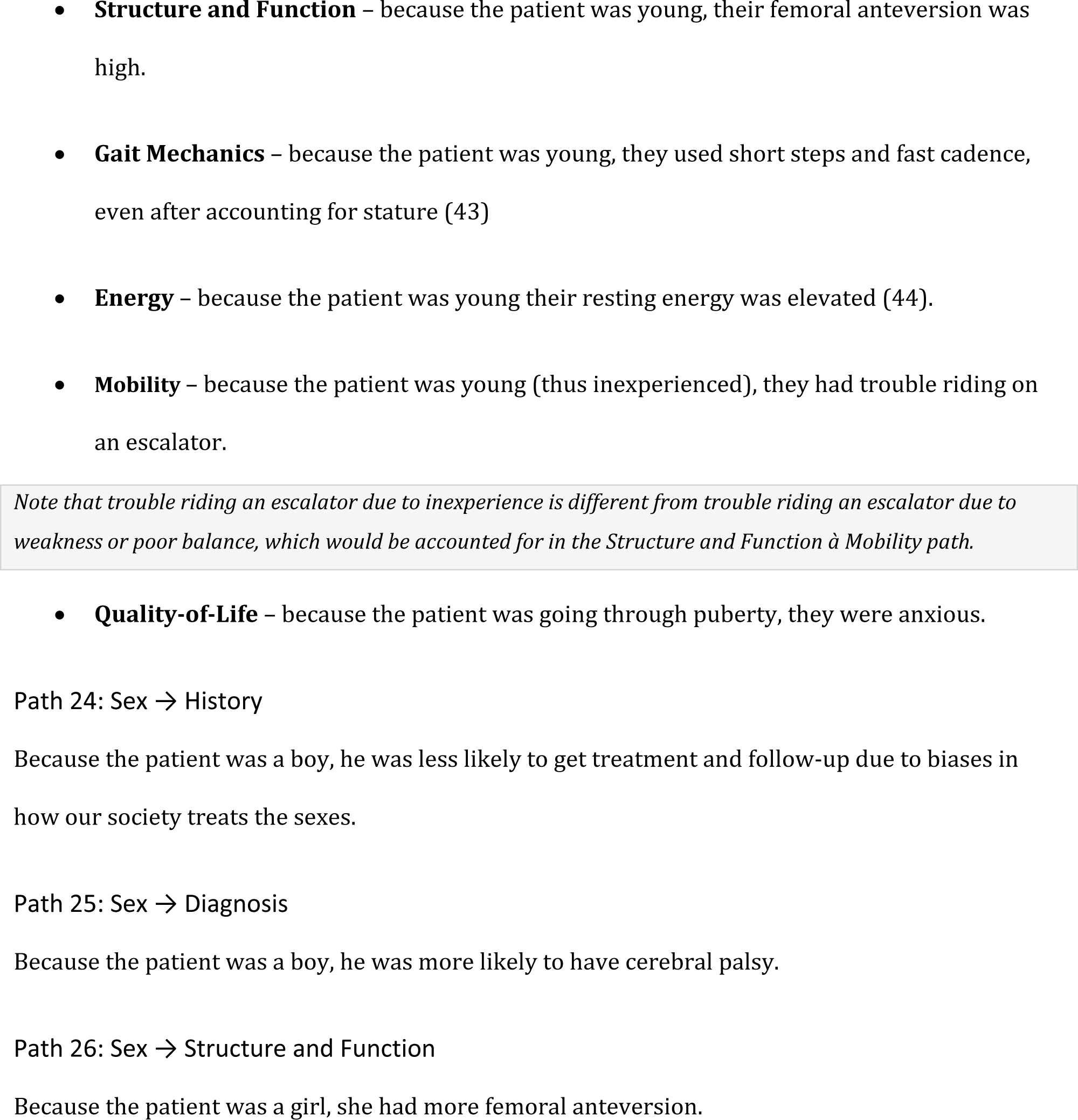

**Appendix 2.**

Variables included in BART Models
| Example | Exposure Node | Outcome Variable | Effect Type | Variables (Adjustment + Exposure) |
| --- | --- | --- | --- | --- |
| 1 | *Structure and Function* | Mean stance foot progression | Total | Lower Extremity Strength, Lower Extremity Static Motor Control, Lower Extremity Spasticity, Dynamic Motor Control, Diagnosis, Age, Max Hip Flex, Max Hip Exten, Max Hip Abd (Knee Extended, Popliteal Angle (Unilateral, Max Knee Flex, Max Knee Exten, Extensor Lag, Max Ankle Dorsiflex (Knee Extended, Max Ankle Plantarflex, Bimalleolar Axis Angle, Forefoot Var/Val, Forefoot Abd/Add, Hindfoot Var/Val, Midfoot Cavus/Planus, Troch Prominence Test Angle |
| 2 | *Structure and Function* | FAQt | Total | Lower Extremity Strength, Lower Extremity Static Motor Control, Lower Extremity Spasticity, Dynamic Motor Control, Diagnosis, Age, Max Hip Flex, Max Hip Exten, Max Hip Abd (Knee Extended, Popliteal Angle (Unilateral, Max Knee Flex, Max Knee Exten, Extensor Lag, Max Ankle Dorsiflex (Knee Extended, Max Ankle Plantarflex, Bimalleolar Axis Angle, Forefoot Var/Val, Forefoot Abd/Add, Hindfoot Var/Val, Midfoot Cavus/Planus, Troch Prominence Test Angle |
| 3 | *Gait Mechanics* | Metabolic Walking Power | Total | Lower Extremity Strength, Lower Extremity Static Motor Control, Lower Extremity Spasticity, Dynamic Motor Control, Diagnosis, Mean Pelvic Tilt, Range-of-Motion Pelvic Tilt, Min. Stance Hip Flexion, Mean Stance Hip Rotation, Init. Contact Knee Flexion, Min. Stance Knee Flexion, Range-of-Motion Knee Flexion, Max. Swing Knee Flexion, Mean Stance Knee Rotation, Init. Contact Ankle Dorsiflexion, Mean Stance Ankle Dorsiflexion, Mean Swing Ankle Dorsiflexion, Mean Stance Foot Progression, Normalized Step Length, Normalized Cadence, Age, Max Hip Flex, Max Hip Exten, Max Hip Abd (Knee Extended, Popliteal Angle (Unilateral, Max Knee Flex, Max Knee Exten, Extensor Lag, Max Ankle Dorsiflex (Knee Extended, Max Ankle Plantarflex, Troch Prominence Test Angle, Bimalleolar Axis Angle, Forefoot Var/Val, Forefoot Abd/Add, Hindfoot Var/Val, Midfoot Cavus/Planus |
| 4 | *Age* | Metabolic Walking Power | Direct | Lower Extremity Strength, Lower Extremity Static Motor Control, Lower Extremity Spasticity, Dynamic Motor Control, Diagnosis, Mean Pelvic Tilt, Range-of-Motion Pelvic Tilt, Min. Stance Hip Flexion, Mean Stance Hip Rotation, Min. Stance Knee Flexion, Init. Contact Knee Flexion, Range-of-Motion Knee Flexion, Max. Swing Knee Flexion, Mean Stance Knee Rotation, Init. Contact Ankle Dorsiflexion, Mean Stance Ankle Dorsiflexion, Mean Swing Ankle Dorsiflexion, Mean Stance Foot Progression, Normalized Step Length, Normalized Cadence, Max Hip Flex, Max Hip Exten, Max Hip Abd (Knee Extended, Popliteal Angle (Unilateral, Max Knee Flex, Max Knee Exten, Extensor Lag, Max Ankle Dorsiflex (Knee Extended, Max Ankle Plantarflex, Troch Prominence Test Angle, Bimalleolar Axis Angle, Forefoot Var/Val, Forefoot Abd/Add, Hindfoot Var/Val, Midfoot Cavus/Planus, Age |
| 5 | *Structure and Function* | Activities of Daily Living and Independence | Total | Lower Extremity Strength, Lower Extremity Static Motor Control, Lower Extremity Spasticity, Dynamic Motor Control, Diagnosis, Age, Max Hip Flex, Max Hip Exten, Max Hip Abd (Knee Extended, Popliteal Angle (Unilateral, Max Knee Flex, Max Knee Exten, Extensor Lag, Max Ankle Dorsiflex (Knee Extended, Max Ankle Plantarflex, Bimalleolar Axis Angle, Forefoot Var/Val, Forefoot Abd/Add, Hindfoot Var/Val, Midfoot Cavus/Planus, Troch Prominence Test Angle |
| 5 | *Gait Mechanics* | Body Image and Self Esteem | Total | Lower Extremity Strength, Lower Extremity Static Motor Control, Lower Extremity Spasticity, Dynamic Motor Control, Diagnosis, Mean Pelvic Tilt, Range-of-Motion Pelvic Tilt, Min. Stance Hip Flexion, Mean Stance Hip Rotation, Init. Contact Knee Flexion, Min. Stance Knee Flexion, Range-of-Motion Knee Flexion, Max. Swing Knee Flexion, Mean Stance Knee Rotation, Init. Contact Ankle Dorsiflexion, Mean Stance Ankle Dorsiflexion, Mean Swing Ankle Dorsiflexion, Mean Stance Foot Progression, Normalized Step Length, Normalized Cadence, Age, Max Hip Flex, Max Hip Exten, Max Hip Abd (Knee Extended, Popliteal Angle (Unilateral, Max Knee Flex, Max Knee Exten, Extensor Lag, Max Ankle Dorsiflex (Knee Extended, Max Ankle Plantarflex, Troch Prominence Test Angle, Bimalleolar Axis Angle, Forefoot Var/Val, Forefoot Abd/Add, Hindfoot Var/Val, Midfoot Cavus/Planus |

